# Risk factors for SARS-CoV-2 infection after primary vaccination with ChAdOx1 nCoV-19 or BNT1262b2 and after booster vaccination with BNT1262b2 or mRNA-1273: a population-based cohort study (COVIDENCE UK)

**DOI:** 10.1101/2022.03.11.22272276

**Authors:** Giulia Vivaldi, David A Jolliffe, Hayley Holt, Florence Tydeman, Mohammad Talaei, Gwyneth A Davies, Ronan A Lyons, Christopher J Griffiths, Frank Kee, Aziz Sheikh, Seif O Shaheen, Adrian R Martineau

## Abstract

**Background:** Little is known about the relative influence of demographic, behavioural, and vaccine-related factors on risk of post-vaccination SARS-CoV-2 infection. We aimed to identify risk factors for SARS-CoV-2 infection after primary and booster vaccinations.

**Methods:** We undertook a prospective population-based study in UK adults (≥16 years) vaccinated against SARS-CoV-2, including data from Jan 12, 2021, to Feb 21, 2022. We modelled risk of post-vaccination SARS-CoV-2 infection separately for participants who had completed a primary course of vaccination (two-dose or, in the immunosuppressed, three-dose course of either ChAdOx1 nCoV-19 [ChAdOx1] or BNT1262b2) and for those who had additionally received a booster dose (BNT1262b2 or mRNA-1273). Cox regression models were used to explore associations between sociodemographic, behavioural, clinical, pharmacological, and nutritional factors and breakthrough infection, defined as a self-reported positive result on a lateral flow or reverse transcription PCR (RT-PCR) test for SARS-CoV-2. Models were further adjusted for weekly SARS-CoV-2 incidence at the local (lower tier local authority) level.

**Findings:** 14,713 participants were included in the post-primary analysis and 10,665 in the post-booster analysis, with a median follow-up of 203 days (IQR 195–216) in the post-primary cohort and 85 days (66–103) in the post-booster cohort. 1051 (7.1%) participants in the post-primary cohort and 1009 (9.4%) participants in the post-booster cohort reported a breakthrough SARS-CoV-2 infection. A primary course of ChAdOx1 (*vs* BNT182b2) was associated with higher risk of infection, both in the post-primary cohort (adjusted hazard ratio 1.63, 95% CI 1.41–1.88) and in the post-booster cohort after boosting with mRNA-1273 (1.29 [1.03–1.61] *vs* BNT162b2 primary plus BNT162b2 booster). A lower risk of breakthrough infection was associated with older age (post-primary: 0.96 [0.96–0.97] per year; post-booster: 0.97 [0.96–0.98]), whereas a higher risk of breakthrough infection was associated with lower levels of education (post-primary: 1.66 [1.35–2.06] for primary or secondary *vs* postgraduate; post-booster: 1.36 [1.08–1.71]) and at least three weekly visits to indoor public places (post-primary: 1.38 [1.15–1.66] *vs* none; post-booster: 1.33 [1.10–1.60]).

**Conclusions:** Vaccine type, socioeconomic status, age, and behaviours affect risk of breakthrough SARS-CoV-2 infection following a primary schedule and a booster dose.

**Research in context:** *Evidence before this study:* We searched PubMed, medRxiv, and Google Scholar for papers published up to Feb 18, 2022, using the search terms (breakthrough OR post-vaccin*) AND (SARS-CoV-2 OR COVID) AND (disease OR infection) AND (determinant OR “risk factor” OR associat*), with no language restrictions. Existing studies on risk factors for breakthrough SARS-CoV-2 infection among vaccinated individuals have found associations with age, comorbidities, vaccine type, and previous infection; however, findings have been inconsistent across studies. Most studies have been limited to specific subgroups or have focused on severe outcomes, and very few have considered breakthrough infections after a booster dose or have adjusted for behaviours affecting exposure to other people.

*Added value of this study:* This study is among the first to provide a detailed analysis of a wide range of risk factors for breakthrough SARS-CoV-2 infection, both after the primary course of vaccination and after a booster dose. Our large study size and detailed data have allowed us to investigate associations with various sociodemographic, clinical, pharmacological, and nutritional factors. Monthly follow-up data have additionally given us the opportunity to consider the effects of behaviours that may have changed across the pandemic, while adjusting for local SARS-CoV-2 incidence.

*Implications of all the available evidence:* Our findings add to growing evidence that risk factors for SARS-CoV-2 infection after primary or booster vaccinations can differ to those in unvaccinated populations, with effects attenuated for previously observed risk factors such as body-mass index and Asian ethnicity. The clear difference we observed between the efficacies of ChAdOx1 and BNT162b2 as the primary course of vaccination appears to have been reduced by the use of BNT162b2 boosters, but not by mNRA-1273 boosters. As more countries introduce booster vaccinations, future population-based studies with longer follow-up will be needed to investigate our findings further.

## Introduction

Vaccination against SARS-CoV-2 has been a key strategy to control the COVID-19 pandemic. With more than 10.7 billion doses administered worldwide, and more than 63% of the world’s population having received at least one dose by March, 2022,^1^ we are moving into the post-vaccination era. Vaccination substantially reduces both COVID-19 disease severity and mortality,^2^ but its effects on transmission appear to be more modest.^3^ Additionally, waning protection^4^ and the emergence of variants with increased transmissibility and immune evasion^5^ increase the likelihood of post-vaccination, or breakthrough, infections.

The picture is further complicated by the introduction of so-called booster doses of SARS-CoV-2 vaccines. The emergence of the omicron variant of concern led to the expedited booster rollout in the UK,^6^ where more than 66% of the population aged 12 years and older had received a booster or third vaccine dose by March, 2022.^7^ The vaccinated population in the UK is now split into two main groups: those who have received a complete primary course and those who have additionally received an (often heterologous) booster dose. As more countries roll out booster programmes,^8^ both groups need to be studied to fully capture their respective risks for breakthrough SARS-CoV-2 infection, and to better understand the effects of booster vaccinations.

While much research has been done on risk factors for SARS-CoV-2 infection in unvaccinated populations, risk factors for breakthrough infection in vaccinated individuals are less well understood and existing studies present conflicting findings. Whereas some studies have reported a higher risk of breakthrough infection among individuals with comorbidities,^9,10^ other large-scale studies have found no associations with pre-existing conditions.^2^ Similarly, several studies have suggested that older age is associated with increased risk of breakthrough infections,^11,12^ whereas others have found no association^13,14^ or an inverse association.^2^ Most studies on risk of breakthrough infection have been limited to specific subgroups,^10,13,15^ focused solely on severe outcomes,^16,17^ or only considered infections before a booster dose.^2,9^ Additionally, the few studies considering behaviours, such as levels of physical activity or journeys on public transport, have relied on baseline values,^2^ and so have been unable to capture changes in behaviour that may have occurred over the various stages of the pandemic.

We therefore did a prospective, nationwide, population-based study in UK adults to investigate the risk factors for breakthrough SARS-CoV-2 infection after primary vaccination and after booster vaccination. We included data on three widely used vaccines and considered a range of potential sociodemographic, clinical, pharmacological, and nutritional determinants of response to vaccination and susceptibility to infection, as well as behavioural factors derived from monthly follow-up questionnaires.

## Methods

### Study design and participants

COVIDENCE UK is a prospective, longitudinal, population-based observational study of COVID-19 in the UK population (www.qmul.ac.uk/covidence). Inclusion criteria were age 16 years or older and UK residence at enrolment, with no exclusion criteria. Participants were invited via a national media campaign to complete an online baseline questionnaire and monthly follow-up questionnaires to capture information on potential symptoms of COVID-19, results of nose or throat swab tests for SARS-CoV-2, COVID-19 vaccination status, and details of a wide range of potential determinants of vaccine response and SARS-CoV-2 exposure. The study was launched on May 1, 2020, and closed to enrolment on Oct 6, 2021. Further details on COVIDENCE UK have been published elsewhere.^18,19^ This analysis is based on monthly follow-up data to Feb 21, 2022.

For the post-primary analysis, we included all participants who had received a two-dose or three-dose primary vaccination course of either the ChAdOx1 nCoV-19 (Oxford–AstraZeneca; hereafter ChAdOx1) or BNT1262b2 mRNA (Pfizer–BioNTech) vaccines. For the booster analysis, we considered all participants who had additionally received a booster dose of BNT1262b2 or mRNA-1273 (Moderna). Participants were considered fully vaccinated 14 days after their second vaccine dose, if receiving a two-dose primary course, or 14 days after their third dose, if receiving a three-dose primary course (offered to immunosuppressed people in the UK^20^). Participants were considered boosted 14 days after their third vaccine dose, if receiving a two-dose primary course, or 14 days after their fourth vaccine dose, if receiving a three-dose primary course.

COVIDENCE UK is registered with ClinicalTrials.gov, NCT04330599, and was approved by Leicester South Research Ethics Committee (ref 20/EM/0117). All participants provided informed consent to participate.

### Outcomes

The primary outcome was incident SARS-CoV-2 after primary or booster vaccinations, defined as a self-reported positive result on a lateral flow or reverse transcription PCR (RT-PCR) test for SARS-CoV-2.

### Independent variables

82 potential determinants of breakthrough infection were chosen a priori for inclusion in our models, covering vaccination type and timing; previous SARS-CoV-2 infection; sociodemographic, occupational, and lifestyle factors; longstanding medical conditions and prescribed medication use; Bacille Calmette Guérin vaccine status; and nutritional factors. All included factors have previously been found to be independently associated with risk of COVID-19 or pre-vaccination or post-vaccination SARS-CoV-2 antibody response.^18,19,21^ Previous infection was defined as reporting a positive result on a lateral flow or RT-PCR test for SARS-CoV-2 before start of the post-vaccination follow-up period.

When considering time-varying factors that could plausibly affect the immune response to vaccination (ie, exercise, sleep, alcohol use, smoking or vaping, anxiety, general health, use of nutritional supplements, and season of vaccination), we focused on the first vaccine dose for the post-primary analysis and on the booster dose for the post-booster analysis. We used the values from the last available monthly questionnaire before the vaccination date of interest (date of first vaccine dose, when considering post-primary outcomes, and date of booster dose, when considering post-booster outcomes). For time-varying covariates taken from monthly questionnaires that could affect SARS-CoV-2 exposure, we included all post-vaccination values observed over the follow-up period.

For post-primary outcomes, the inter-dose interval between vaccinations was calculated as the time between the first and second doses received, regardless of whether the participant was receiving a two-dose or three-dose regimen. For the post-booster analysis, the inter-dose interval was calculated as the time between the last primary course vaccination received and the booster vaccination.

To produce participant-level covariates for each class of medications investigated, questionnaire responses were mapped to drug classes listed in the British National Formulary or the DrugBank and Electronic Medicines Compendium databases if not explicitly listed in the British National Formulary, as previously described.^18^ Participants were assigned Index of Multiple Deprivation (IMD) 2019 scores, or equivalent scores for devolved administrations, according to their postcode.

### Statistical analysis

We carried out two separate analyses: a post-primary analysis and a post-booster analysis. Participants were included in the post-primary analysis when fully vaccinated, and were excluded from the analysis either at time of breakthrough infection, 13 days after their booster dose, or at the end of follow-up, whichever came first. Participants were included in the post-booster analysis 14 days after their booster dose, and were excluded from the analysis either at time of breakthrough infection or end of follow-up, whichever came first. Participants who experienced a breakthrough infection in the post-primary analysis were classified as having had a previous SARS-CoV-2 infection upon entry into the post-booster analysis.

We used Cox proportional hazards models to estimate the hazard ratios (HRs) for potential determinants of breakthrough infection after vaccination. We first estimated HRs in minimally adjusted models (adjusted for age and sex), and included all factors independently associated with breakthrough infection at the 10% significance level in fully adjusted models. We additionally adjusted for the local weekly SARS-CoV-2 incidence, using incidence rates reported for each lower tier local authority^7^ and assigning them to participants according to their residential postcode. For participants without postcode data, we assigned them the weekly incidence of their country of residence, if available, or of the UK as a whole. We replaced missing values of time-varying covariates with mean values calculated for each participant over their follow-up period; if unavailable, we used the last value observed before vaccination.

We used orthogonal polynomial contrasts to test for linear trends in ordinal variables included in fully adjusted models. Correlation matrices were examined to check for collinearity between variables. The proportional hazards assumption for each model was tested using Schoenfeld residuals and visual assessment of log–log plots of survival.

Analyses were done using Stata (version 17.0).

## Results

14,713 participants were included in the post-primary analysis and 10,665 were included in the post-booster analysis (figure 1), with a median follow-up of 203 days (IQR 195–216) in the post-primary cohort and 85 days (66–103) in the post-booster cohort (table 1). The cohort characteristics were largely similar, with boosted participants being slightly older (table 1). The earliest entry into the post-primary cohort was Jan 12, 2021, and the earliest entry into the post-booster cohort was Sept 5, 2021.

**Table 1:** Participant characteristics.

|  | Post-primary cohort<br>(n=14,713) | Post-booster cohort<br>(n=10,665) |
| --- | --- | --- |
| **Sociodemographic and lifestyle factors** |  |  |
| Age, years | 62.9 (54.3–69.5) | 64.3 (56.8–70.1) |
| <30 | 324 (2.2%) | 124 (1.2%) |
| 30 to <40 | 693 (4.7%) | 328 (3.1%) |
| 40 to <50 | 1464 (10.0%) | 823 (7.7%) |
| 50 to <60 | 3469 (23.6%) | 2376 (22.3%) |
| 60 to <70 | 5342 (36.3%) | 4311 (40.4%) |
| ≥70 | 3421 (23.3%) | 2703 (25.3%) |
| Sex |  |  |
| Female | 10,430 (70.9%) | 7511 (70.4%) |
| Male | 4283 (29.1%) | 3154 (29.6%) |
| Ethnicity* |  |  |
| White | 14,068 (95.6%) | 10,290 (96.5%) |
| Mixed, multiple, or other ethnic groups | 355 (2.4%) | 224 (2.1%) |
| South Asian | 212 (1.4%) | 114 (1.1%) |
| Black, African, Caribbean, or Black British | 77 (0.5%) | 36 (0.3%) |
| Country of residence* |  |  |
| England | 13021 (88.8%) | 9515 (89.3%) |
| Northern Ireland | 253 (1.7%) | 167 (1.6%) |
| Scotland | 850 (5.8%) | 616 (5.8%) |
| Wales | 535 (3.6%) | 363 (3.4%) |
| Housing* |  |  |
| Owns own home | 9527 (64.8%) | 7535 (70.7%) |
| Mortgage | 3418 (23.2%) | 2157 (20.2%) |
| Privately renting | 866 (5.9%) | 489 (4.6%) |
| Renting from council | 446 (3.0%) | 253 (2.4%) |
| Other | 452 (3.1%) | 229 (2.1%) |
| Number of people per bedroom* |  |  |
| <1 | 10,089 (69.0%) | 7724 (72.9%) |
| 1 to <2 | 4252 (29.1%) | 2719 (25.7%) |
| ≥2 | 272 (1.9%) | 152 (1.4%) |
| Quartiles of IMD rank* |  |  |
| Q4 (least deprived) | 3942 (26.9%) | 3000 (28.2%) |
| Q3 | 3789 (25.9%) | 2838 (26.6%) |
| Q2 | 3635 (24.8%) | 2577 (24.2%) |
| Q1 (most deprived) | 3282 (22.4%) | 2238 (21.0%) |
| Frontline worker* |  |  |
| No | 11,974 (81.5%) | 8875 (83.3%) |
| Non-health or care | 1464 (10.0%) | 924 (8.7%) |
| Health or care | 1261 (8.6%) | 857 (8.0%) |
| Highest educational level attained* |  |  |
| Primary or secondary | 1562 (10.6%) | 1090 (10.2%) |
| Higher or further (A levels) | 2123 (14.4%) | 1541 (14.5%) |
| College or university | 6535 (44.5%) | 4767 (44.7%) |
| Post-grad | 4477 (30.5%) | 3256 (30.6%) |
| Smoking status |  |  |
| Never-smoker | 8310 (56.5%) | 6111 (57.3%) |
| Ex-smoker | 5708 (38.8%) | 4128 (38.7%) |
| Current smoker | 695 (4.7%) | 426 (4.0%) |
| Vaping status* |  |  |
| Never-vaper | 13,823 (94.2%) | 10,131 (95.2%) |
| Ex-vaper | 435 (3.0%) | 265 (2.5%) |
| Current vaper | 414 (2.8%) | 242 (2.3%) |
| Alcohol consumption, units per week |  |  |
| 0 | 4143 (28.2%) | 2833 (26.6%) |
| 1–7 | 5168 (35.1%) | 3768 (35.3%) |
| 8–14 | 2902 (19.7%) | 2192 (20.6%) |
| ≥15 | 2500 (17.0%) | 1872 (17.6%) |
| **Vaccination and breakthrough infections** |  |  |
| Primary vaccination course |  |  |
| ChAdOx1 | 8969 (61.0%) | 6537 (61.3%) |
| BNT162b2 | 5744 (39.0%) | 4128 (38.7%) |
| Three-dose regimen | 71 (0.4%) | 31 (0.3%) |
| Inter-dose interval (first to second dose), weeks |  |  |
| <6 | 482 (3.3%) | 345 (3.2%) |
| 6–10 | 5398 (36.7%) | 3774 (35.4%) |
| >10 | 8833 (60.0%) | 6546 (61.4%) |
| Booster vaccine combination |  |  |
| ChAdOx1 primary plus BNT162b2 booster | .. | 5288 (48.8%) |
| ChAdOx1 primary plus mRNA-1273 booster | .. | 1364 (12.6%) |
| BNT162b2 primary plus BNT162b2 booster | .. | 1825 (35.3%) |
| BNT162b2 primary plus mRNA-1273 booster | .. | 371 (3.4%) |
| Inter-dose interval (primary to booster), weeks |  |  |
| <20 | .. | 79 (0.7%) |
| 20–30 | .. | 9706 (91.0%) |
| >30 | .. | 880 (8.3%) |
| Days of follow-up | 203 (195–216) | 85 (66–103) |
| Breakthrough infection | 1051 (7.1%) | 1005 (9.4%) |
| Hospitalised | 10 (1.0%) | 2 (0.2%) |
| Days to breakthrough infection | 160 (115–196) | 68 (48–89) |
| **Medical conditions** |  |  |
| BMI, kg/m<sup>2</sup>* |  |  |
| <25 | 7112 (48.4%) | 5229 (49.1%) |
| 25 to <30 | 4747 (32.3%) | 3469 (32.6%) |
| ≥30 | 2830 (19.3%) | 1952 (18.3%) |
| Arterial disease | 810 (5.5%) | 614 (5.8%) |
| Asthma | 2464 (16.7%) | 1726 (16.2%) |
| Atopy | 3765 (25.6%) | 2795 (26.2%) |
| Autoimmune disease | 1361 (9.3%) | 962 (9.0%) |
| Cancer |  |  |
| Past (cured or in remission) | 1334 (9.1%) | 1044 (9.8%) |
| Present (active treatment) | 134 (0.9%) | 102 (1.0%) |
| COPD | 328 (2.2%) | 234 (2.2%) |
| Diabetes type |  |  |
| Pre-diabetes | 480 (3.3%) | 356 (3.3%) |
| Diabetes | 749 (5.1%) | 545 (5.1%) |
| Heart disease | 599 (4.1%) | 457 (4.3%) |
| Hypertension | 3359 (22.8%) | 2560 (24.0%) |
| Immunodeficiency | 95 (0.6%) | 67 (0.6%) |
| Kidney disease | 313 (2.1%) | 248 (2.3%) |
| Major neurological conditions | 420 (2.9%) | 313 (2.9%) |

Data are n (%), n/N (%), or median (IQR). BMI = body-mass index. ChAdOx1 = ChAdOx1 nCoV-19. COPD = chronic obstructive pulmonary disease. \*Missing data for ethnicity (post-primary: n=1; post-booster: n=1), country (n=54; n=4), housing (n=4; n=2), number of people per bedroom (n=100; n=70), IMD (n=65; n=12), frontline worker status (n=14; n=9), education (n=16; n=11), vaping (n=41; n=27), and BMI (n=24; n=15).

**Figure 1:**
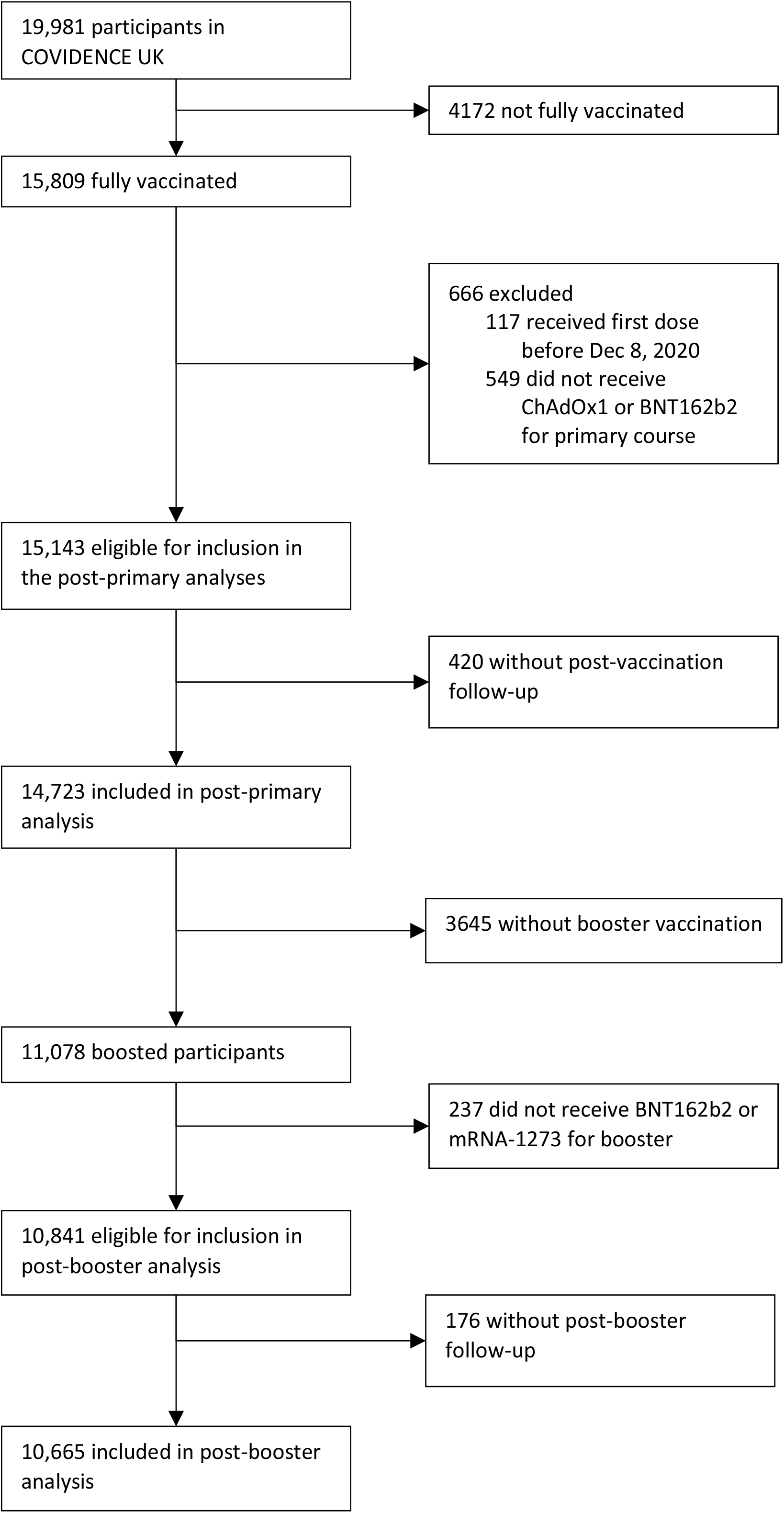
Study flow diagram. ChAdOx1 = ChAdOx1 nCoV-19.

Between Jan 12, 2021, and Feb 21, 2022, 1051 (7.1%) breakthrough infections were reported among fully vaccinated participants. After adjustment for age and sex, 24 factors were associated with risk of post-primary breakthrough infection (table 2; see appendix table S1 for factors for which no association was found). When included together in a fully adjusted model, we observed that primary or secondary education (*vs* postgraduate), household overcrowding (*vs* <1 person per bedroom), sharing a home with schoolchildren, pre-vaccination vaping, any visits to or from other households, frequent visits to indoor public places other than shops (*vs* no visits), greater interval between vaccine doses, a primary course of ChAdOx1 (*vs* BNT162b2), and receiving the first vaccine dose between mid-April and mid-October, 2021, were independently associated with increased risk of breakthrough infection (table 2; figure 2). Older age, being a frontline worker within health or social care (*vs* no frontline occupation), having previously tested positive for SARS-CoV-2, and use of anticholinergic medication were independently associated with reduced risk of breakthrough infection (table 2; figure 2).

**Table 2:**
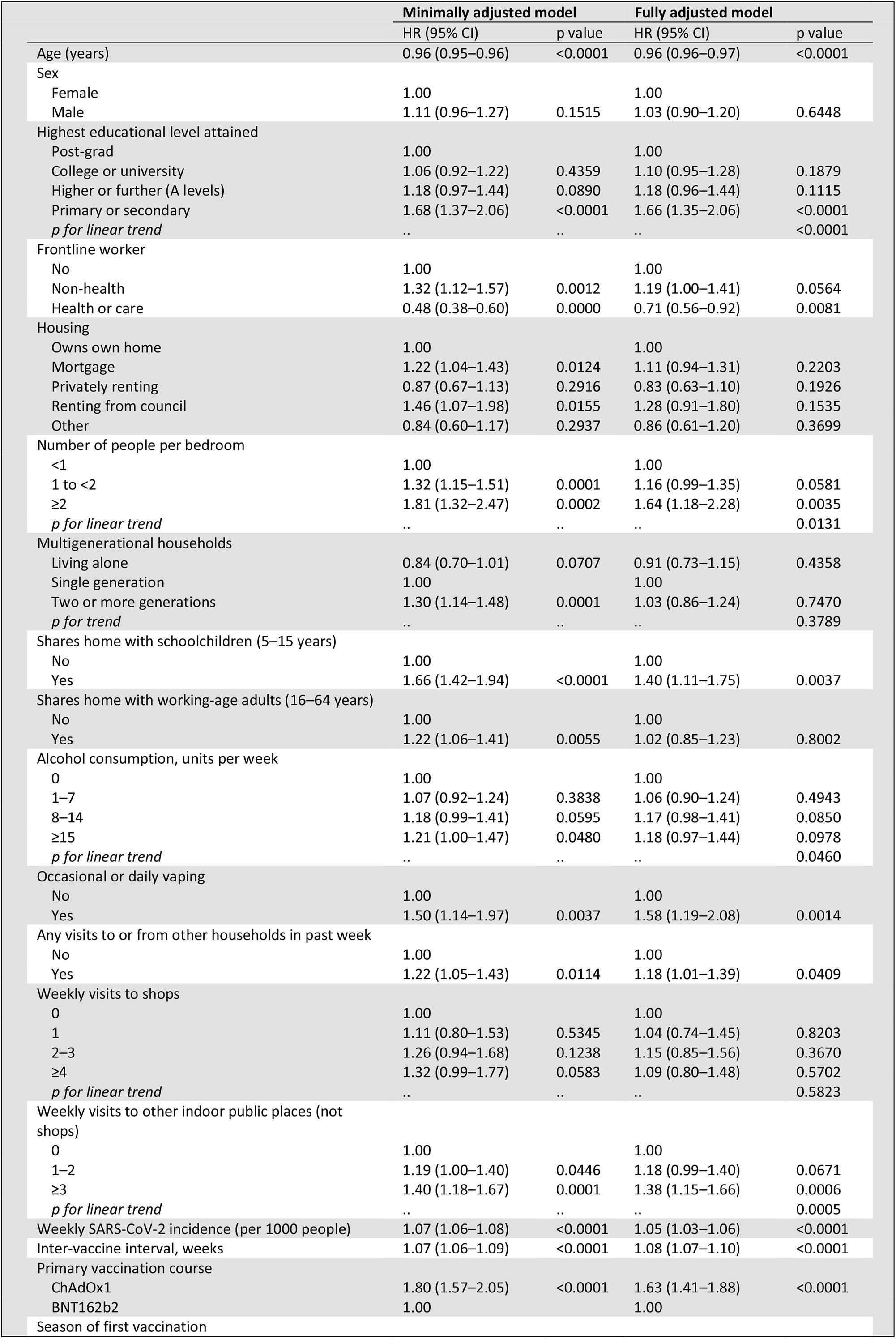
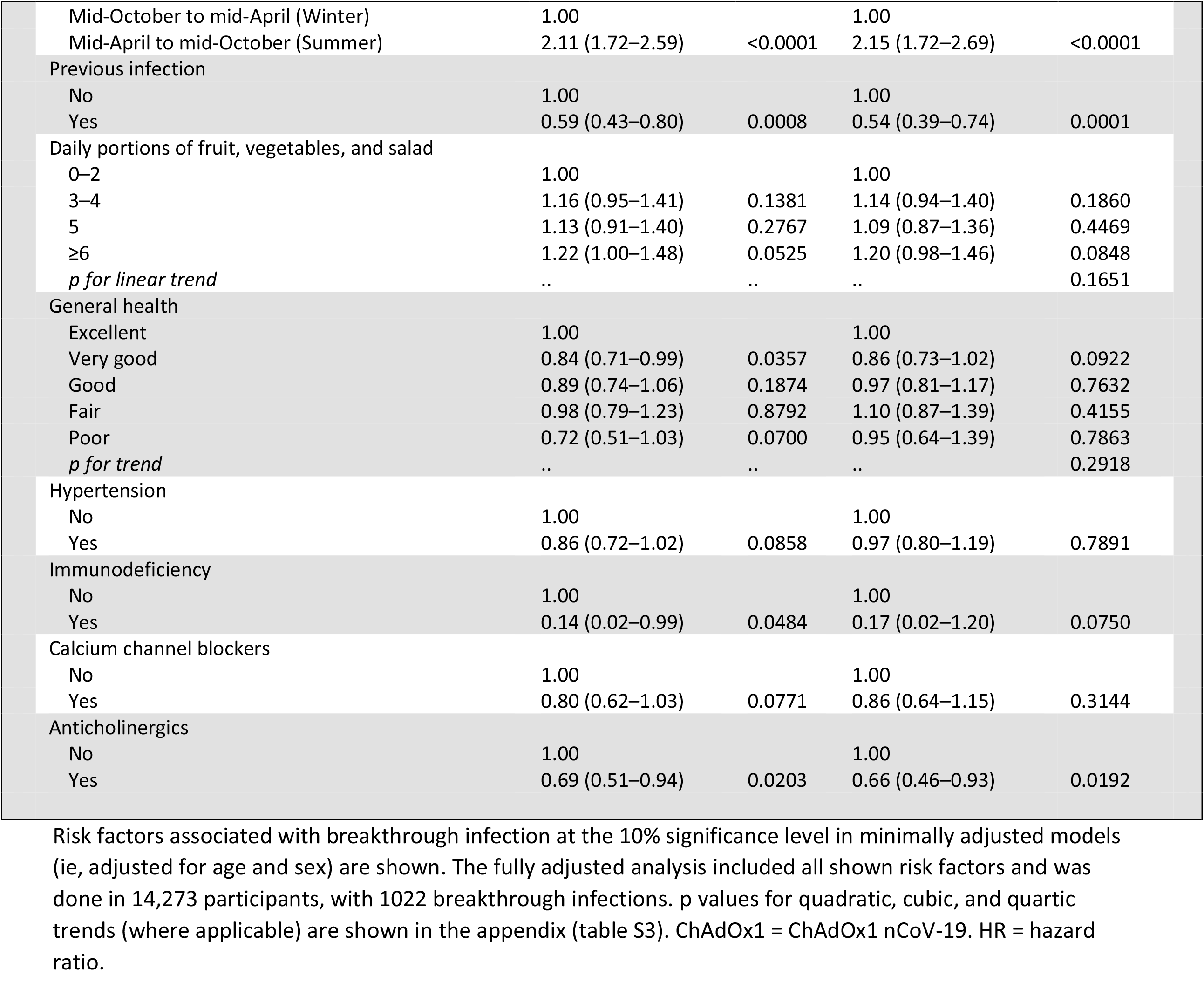
Risk factors for breakthrough SARS-CoV-2 infection in the post-primary cohort.

|  | Minimally adjusted model |  | Fully adjusted model |  |
| --- | --- | --- | --- | --- |
|  | HR (95% CI) | p value | HR (95% CI) | p value |
| Age (years) | 0.96 (0.95–0.96) | <0.0001 | 0.96 (0.96–0.97) | <0.0001 |
| Sex |  |  |  |  |
| Female | 1.00 |  | 1.00 |  |
| Male | 1.11 (0.96–1.27) | 0.1515 | 1.03 (0.90–1.20) | 0.6448 |
| Highest educational level attained |  |  |  |  |
| Post-grad | 1.00 |  | 1.00 |  |
| College or university | 1.06 (0.92–1.22) | 0.4359 | 1.10 (0.95–1.28) | 0.1879 |
| Higher or further (A levels) | 1.18 (0.97–1.44) | 0.0890 | 1.18 (0.96–1.44) | 0.1115 |
| Primary or secondary | 1.68 (1.37–2.06) | <0.0001 | 1.66 (1.35–2.06) | <0.0001 |
| *p for linear trend* | .. | .. | .. | <0.0001 |
| Frontline worker |  |  |  |  |
| No | 1.00 |  | 1.00 |  |
| Non-health | 1.32 (1.12–1.57) | 0.0012 | 1.19 (1.00–1.41) | 0.0564 |
| Health or care | 0.48 (0.38–0.60) | 0.0000 | 0.71 (0.56–0.92) | 0.0081 |
| Housing |  |  |  |  |
| Owns own home | 1.00 |  | 1.00 |  |
| Mortgage | 1.22 (1.04–1.43) | 0.0124 | 1.11 (0.94–1.31) | 0.2203 |
| Privately renting | 0.87 (0.67–1.13) | 0.2916 | 0.83 (0.63–1.10) | 0.1926 |
| Renting from council | 1.46 (1.07–1.98) | 0.0155 | 1.28 (0.91–1.80) | 0.1535 |
| Other | 0.84 (0.60–1.17) | 0.2937 | 0.86 (0.61–1.20) | 0.3699 |
| Number of people per bedroom |  |  |  |  |
| <1 | 1.00 |  | 1.00 |  |
| 1 to <2 | 1.32 (1.15–1.51) | 0.0001 | 1.16 (0.99–1.35) | 0.0581 |
| ≥2 | 1.81 (1.32–2.47) | 0.0002 | 1.64 (1.18–2.28) | 0.0035 |
| *p for linear trend* | .. | .. | .. | 0.0131 |
| Multigenerational households |  |  |  |  |
| Living alone | 0.84 (0.70–1.01) | 0.0707 | 0.91 (0.73–1.15) | 0.4358 |
| Single generation | 1.00 |  | 1.00 |  |
| Two or more generations | 1.30 (1.14–1.48) | 0.0001 | 1.03 (0.86–1.24) | 0.7470 |
| *p for trend* | .. | .. | .. | 0.3789 |
| Shares home with schoolchildren (5–15 years) |  |  |  |  |
| No | 1.00 |  | 1.00 |  |
| Yes | 1.66 (1.42–1.94) | <0.0001 | 1.40 (1.11–1.75) | 0.0037 |
| Shares home with working-age adults (16–64 years) |  |  |  |  |
| No | 1.00 |  | 1.00 |  |
| Yes | 1.22 (1.06–1.41) | 0.0055 | 1.02 (0.85–1.23) | 0.8002 |
| Alcohol consumption, units per week |  |  |  |  |
| 0 | 1.00 |  | 1.00 |  |
| 1–7 | 1.07 (0.92–1.24) | 0.3838 | 1.06 (0.90–1.24) | 0.4943 |
| 8–14 | 1.18 (0.99–1.41) | 0.0595 | 1.17 (0.98–1.41) | 0.0850 |
| ≥15 | 1.21 (1.00–1.47) | 0.0480 | 1.18 (0.97–1.44) | 0.0978 |
| *p for linear trend* | .. | .. | .. | 0.0460 |
| Occasional or daily vaping |  |  |  |  |
| No | 1.00 |  | 1.00 |  |
| Yes | 1.50 (1.14–1.97) | 0.0037 | 1.58 (1.19–2.08) | 0.0014 |
| Any visits to or from other households in past week |  |  |  |  |
| No | 1.00 |  | 1.00 |  |
| Yes | 1.22 (1.05–1.43) | 0.0114 | 1.18 (1.01–1.39) | 0.0409 |
| Weekly visits to shops |  |  |  |  |
| 0 | 1.00 |  | 1.00 |  |
| 1 | 1.11 (0.80–1.53) | 0.5345 | 1.04 (0.74–1.45) | 0.8203 |
| 2–3 | 1.26 (0.94–1.68) | 0.1238 | 1.15 (0.85–1.56) | 0.3670 |
| ≥4 | 1.32 (0.99–1.77) | 0.0583 | 1.09 (0.80–1.48) | 0.5702 |
| *p for linear trend* | .. | .. | .. | 0.5823 |
| Weekly visits to other indoor public places (not shops) |  |  |  |  |
| 0 | 1.00 |  | 1.00 |  |
| 1–2 | 1.19 (1.00–1.40) | 0.0446 | 1.18 (0.99–1.40) | 0.0671 |
| ≥3 | 1.40 (1.18–1.67) | 0.0001 | 1.38 (1.15–1.66) | 0.0006 |
| *p for linear trend* | .. | .. | .. | 0.0005 |
| Weekly SARS-CoV-2 incidence (per 1000 people) | 1.07 (1.06–1.08) | <0.0001 | 1.05 (1.03–1.06) | <0.0001 |
| Inter-vaccine interval, weeks | 1.07 (1.06–1.09) | <0.0001 | 1.08 (1.07–1.10) | <0.0001 |
| Primary vaccination course |  |  |  |  |
| ChAdOx1 | 1.80 (1.57–2.05) | <0.0001 | 1.63 (1.41–1.88) | <0.0001 |
| BNT162b2 | 1.00 |  | 1.00 |  |
| Season of first vaccination |  |  |  |  |
| Mid-October to mid-April (Winter) | 1.00 |  | 1.00 |  |
| Mid-April to mid-October (Summer) | 2.11 (1.72–2.59) | <0.0001 | 2.15 (1.72–2.69) | <0.0001 |
| Previous infection |  |  |  |  |
| No | 1.00 |  | 1.00 |  |
| Yes | 0.59 (0.43–0.80) | 0.0008 | 0.54 (0.39–0.74) | 0.0001 |
| Daily portions of fruit, vegetables, and salad |  |  |  |  |
| 0–2 | 1.00 |  | 1.00 |  |
| 3–4 | 1.16 (0.95–1.41) | 0.1381 | 1.14 (0.94–1.40) | 0.1860 |
| 5 | 1.13 (0.91–1.40) | 0.2767 | 1.09 (0.87–1.36) | 0.4469 |
| ≥6 | 1.22 (1.00–1.48) | 0.0525 | 1.20 (0.98–1.46) | 0.0848 |
| *p for linear trend* | .. | .. | .. | 0.1651 |
| General health |  |  |  |  |
| Excellent | 1.00 |  | 1.00 |  |
| Very good | 0.84 (0.71–0.99) | 0.0357 | 0.86 (0.73–1.02) | 0.0922 |
| Good | 0.89 (0.74–1.06) | 0.1874 | 0.97 (0.81–1.17) | 0.7632 |
| Fair | 0.98 (0.79–1.23) | 0.8792 | 1.10 (0.87–1.39) | 0.4155 |
| Poor | 0.72 (0.51–1.03) | 0.0700 | 0.95 (0.64–1.39) | 0.7863 |
| *p for trend* | .. | .. | .. | 0.2918 |
| Hypertension |  |  |  |  |
| No | 1.00 |  | 1.00 |  |
| Yes | 0.86 (0.72–1.02) | 0.0858 | 0.97 (0.80–1.19) | 0.7891 |
| Immunodeficiency |  |  |  |  |
| No | 1.00 |  | 1.00 |  |
| Yes | 0.14 (0.02–0.99) | 0.0484 | 0.17 (0.02–1.20) | 0.0750 |
| Calcium channel blockers |  |  |  |  |
| No | 1.00 |  | 1.00 |  |
| Yes | 0.80 (0.62–1.03) | 0.0771 | 0.86 (0.64–1.15) | 0.3144 |
| Anticholinergics |  |  |  |  |
| No | 1.00 |  | 1.00 |  |
| Yes | 0.69 (0.51–0.94) | 0.0203 | 0.66 (0.46–0.93) | 0.0192 |

Risk factors associated with breakthrough infection at the 10% significance level in minimally adjusted models (ie, adjusted for age and sex) are shown. The fully adjusted analysis included all shown risk factors and was done in 14,273 participants, with 1022 breakthrough infections. p values for quadratic, cubic, and quartic trends (where applicable) are shown in the appendix (table S3). ChAdOx1 = ChAdOx1 nCoV-19. HR = hazard ratio.

**Figure 2:**
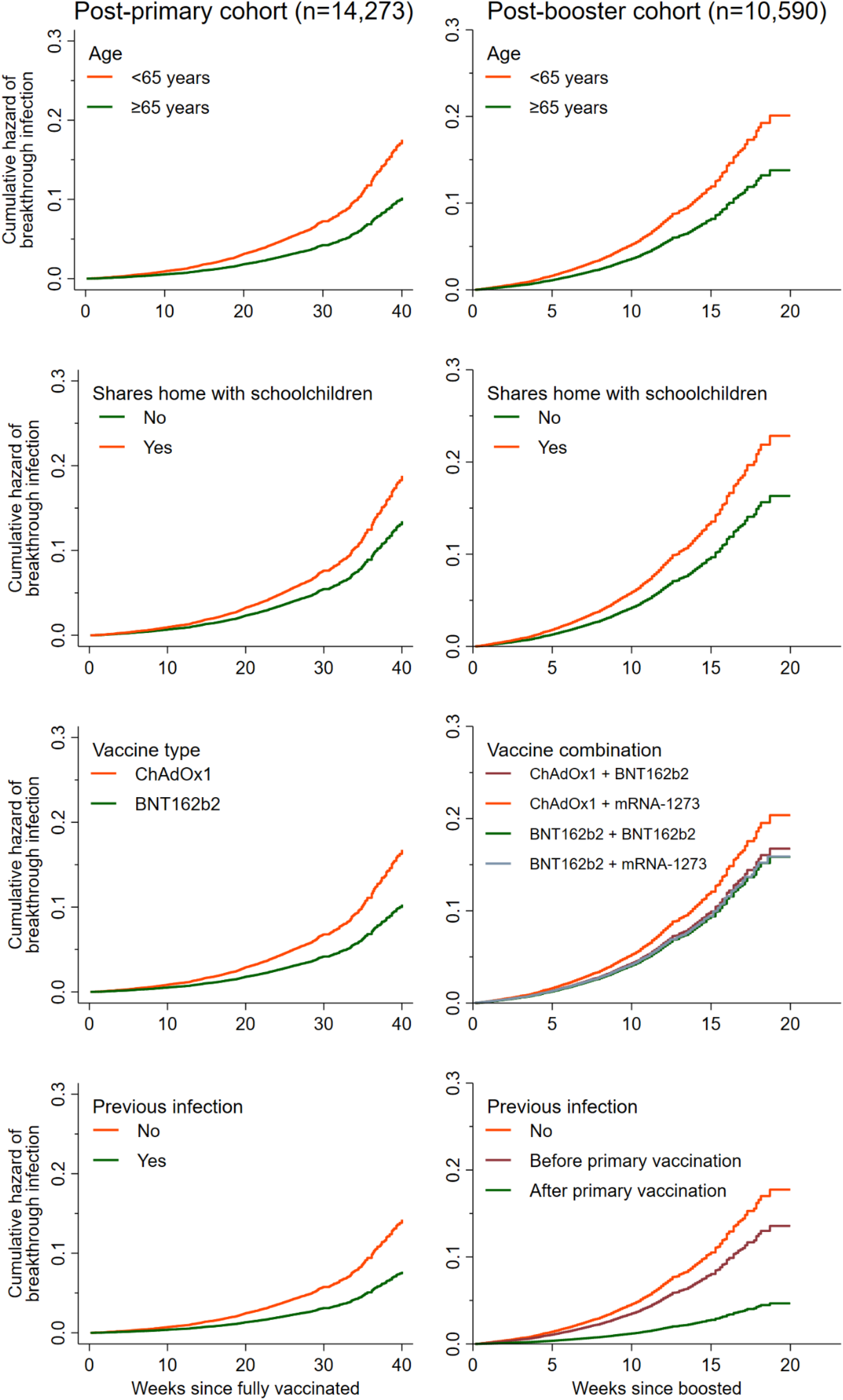
Cumulative hazard of breakthrough infection according to age, sharing a home with schoolchildren, vaccine type, and previous infection for the post-primary and post-booster cohorts. Cumulative hazards shown are from the fully adjusted models. ChAdOx1 = ChAdOx1 nCoV-19.

Between Sept 5, 2021, and Feb 21, 2022, 1009 (9.4%) breakthrough infections were reported among boosted participants, the majority (938 [93.0%]) of which occurred after the omicron variant became dominant in the UK (Dec 17, 2021). After adjustment for age and sex, 31 factors were associated with risk of post-booster breakthrough infection (table 3; see appendix table S2 for factors for which no association was found). When included together in a fully adjusted model, we observed that lower levels of education (*vs* postgraduate), sharing a home with schoolchildren or working-age adults, higher levels of pre-vaccination alcohol consumption (*vs* none), frequent visits to indoor public places other than shops (*vs* none), and a primary course of ChAdOx1 followed by a mRNA-1273 booster (*vs* BNT162b2 for both primary course and booster) were associated with increased risk of breakthrough infection, whereas older age, male sex, mixed or south Asian ethnicity (*vs* White), an average of no more than 5 hours of sleep pre-vaccination (*vs* 7 hours), receiving the booster vaccination between mid-April and mid-October, having had a previous infection after the primary course of vaccination, and taking thiazides were associated with reduced risk (table 3; figure 2). Increased local weekly SARS-CoV-2 incidence was associated with increased risk in all models (tables 2, 3).

**Table 3:** Risk factors for breakthrough SARS-CoV-2 infection in the post-booster cohort.

|  | Minimally adjusted |  | Fully adjusted |  |
| --- | --- | --- | --- | --- |
|  | HR (95% CI) | p value | HR (95% CI) | p value |
| Age, years | 0.96 (0.96–0.97) | <0.0001 | 0.97 (0.96–0.98) | <0.0001 |
| Sex |  |  |  |  |
| Female | 1.00 |  | 1.00 |  |
| Male | 0.93 (0.80–1.07) | 0.3218 | 0.85 (0.73–0.99) | 0.0406 |
| Ethnicity |  |  |  |  |
| White | 1.00 |  | 1.00 |  |
| Mixed, multiple, or other ethnic groups | 0.27 (0.13–0.54) | 0.0002 | 0.28 (0.14–0.57) | 0.0004 |
| South Asian | 0.40 (0.19–0.85) | 0.0174 | 0.44 (0.21–0.92) | 0.0304 |
| Black, African, Caribbean, or Black British | 0.98 (0.37–2.62) | 0.9699 | 0.90 (0.33–2.41) | 0.8285 |
| Highest educational level attained |  |  |  |  |
| Post-grad | 1.00 |  | 1.00 |  |
| College or university | 1.06 (0.92–1.23) | 0.4096 | 1.07 (0.92–1.24) | 0.4019 |
| Higher or further (A levels) | 1.19 (0.98–1.45) | 0.0805 | 1.21 (0.99–1.48) | 0.0598 |
| Primary or secondary | 1.30 (1.04–1.63) | 0.0210 | 1.36 (1.08–1.71) | 0.0092 |
| *p for trend* | .. | .. | .. | 0.0043 |
| Frontline worker |  |  |  |  |
| No | 1.00 |  | 1.00 |  |
| Non-health | 1.20 (0.98–1.47) | 0.0823 | 1.11 (0.90–1.37) | 0.3276 |
| Health or care | 0.74 (0.60–0.91) | 0.0043 | 0.91 (0.71–1.16) | 0.4294 |
| Housing |  |  |  |  |
| Owns own home | 1.00 |  | 1.00 |  |
| Mortgage | 1.23 (1.04–1.45) | 0.0137 | 1.20 (1.01–1.42) | 0.0379 |
| Privately renting | 1.27 (0.97–1.66) | 0.0864 | 1.38 (1.04–1.83) | 0.0236 |
| Renting from council | 0.93 (0.60–1.44) | 0.7454 | 1.08 (0.69–1.70) | 0.7223 |
| Other | 0.92 (0.60–1.39) | 0.6819 | 1.04 (0.68–1.59) | 0.8728 |
| Multigenerational households |  |  |  |  |
| Living alone | 0.78 (0.65–0.94) | 0.0080 | 0.92 (0.74–1.13) | 0.4258 |
| Single generation | 1.00 |  | 1.00 |  |
| Two or more generations | 1.15 (0.99–1.32) | 0.0615 | 0.98 (0.82–1.17) | 0.7882 |
| *p for trend* | .. | .. | .. | 0.6556 |
| Shares home with schoolchildren (5–15 years) |  |  |  |  |
| No | 1.00 |  | 1.00 |  |
| Yes | 1.45 (1.19–1.76) | 0.0002 | 1.40 (1.09–1.79) | 0.0080 |
| Shares home with working-age adults (16–64 years) |  |  |  |  |
| No | 1.00 |  | 1.00 |  |
| Yes | 1.38 (1.19–1.59) | <0.0001 | 1.28 (1.08–1.52) | 0.0051 |
| Alcohol consumption, units per week |  |  |  |  |
| 0 | 1.00 |  | 1.00 |  |
| 1–7 | 1.19 (1.01–1.40) | 0.0391 | 1.06 (0.90–1.26) | 0.4853 |
| 8–14 | 1.21 (1.00–1.45) | 0.0502 | 1.02 (0.84–1.23) | 0.8692 |
| ≥15 | 1.53 (1.27–1.83) | <0.0001 | 1.30 (1.08–1.58) | 0.0067 |
| *p for trend* | .. | .. | .. | 0.0234 |
| Travelled outside of the UK since last questionnaire |  |  |  |  |
| No | 1.00 |  | 1.00 |  |
| Yes | 1.24 (1.06–1.44) | 0.0073 | 1.11 (0.94–1.30) | 0.2153 |
| Weekly journeys on public transport |  |  |  |  |
| 0 | 1.00 |  | 1.00 |  |
| 1–5 | 1.09 (0.94–1.28) | 0.2516 | 1.00 (0.85–1.17) | 0.9752 |
| ≥6 | 1.27 (1.02–1.58) | 0.0346 | 1.15 (0.91–1.46) | 0.2288 |
| *p for trend* | .. | .. | .. | 0.4084 |
| Any visits to or from other households in past week |  |  |  |  |
| No | 1.00 |  | 1.00 |  |
| Yes | 1.16 (0.98–1.36) | 0.0837 | 1.10 (0.93–1.29) | 0.2802 |
| Weekly visits to shops |  |  |  |  |
| 0 | 1.00 |  | 1.00 |  |
| 1 | 0.94 (0.68–1.31) | 0.7216 | 0.94 (0.67–1.31) | 0.7020 |
| 2–3 | 1.12 (0.84–1.49) | 0.4335 | 1.06 (0.79–1.43) | 0.6921 |
| ≥4 | 1.40 (1.06–1.85) | 0.0196 | 1.18 (0.87–1.59) | 0.2815 |
| *p for trend* | .. | .. | .. | 0.0418 |
| Weekly visits to other indoor public places (not shops) |  |  |  |  |
| 0 | 1.00 |  | 1.00 |  |
| 1–2 | 1.18 (0.99–1.40) | 0.0663 | 1.09 (0.92–1.31) | 0.3219 |
| ≥3 | 1.59 (1.33–1.89) | <0.0001 | 1.33 (1.10–1.60) | 0.0035 |
| *p for trend* | .. | .. | .. | 0.0021 |
| Vigorous physical exercise, h per week |  |  |  |  |
| 0 | 0.84 (0.71–0.99) | 0.0373 | 0.89 (0.75–1.06) | 0.1872 |
| 1 | 0.92 (0.78–1.07) | 0.2671 | 0.91 (0.78–1.07) | 0.2621 |
| 2 | 1.00 |  | 1.00 |  |
| *p for trend* | .. | .. | .. | 0.2058 |
| Actual sleep, h/night |  |  |  |  |
| ≤5 | 0.71 (0.56–0.90) | 0.0050 | 0.73 (0.57–0.93) | 0.0100 |
| 6 | 1.01 (0.87–1.17) | 0.9218 | 1.01 (0.87–1.18) | 0.8513 |
| 7 | 1.00 |  | 1.00 |  |
| ≥8 | 0.98 (0.83–1.16) | 0.7998 | 1.01 (0.85–1.20) | 0.8921 |
| *p for trend* | .. | .. | .. | 0.0547 |
| Weekly SARS-CoV-2 incidence | 1.09 (1.08–1.10) | <0.0001 | 1.09 (1.08–1.10) | <0.0001 |
| Inter-vaccine interval (primary to booster), weeks | 0.97 (0.95–0.99) | 0.0077 | 0.98 (0.96–1.01) | 0.1867 |
| Combination of primary and booster vaccinations |  |  |  |  |
| ChAdOx1 plus BNT162b2 booster | 1.16 (1.01–1.33) | 0.0341 | 1.06 (0.91–1.23) | 0.4755 |
| ChAdOx1 plus mRNA-1273 booster | 1.41 (1.14–1.74) | 0.0014 | 1.29 (1.03–1.61) | 0.0284 |
| BNT162b2 plus BNT162b2 booster | 1.00 |  | 1.00 |  |
| BNT162b2 plus mRNA-1273 booster | 1.07 (0.73–1.56) | 0.7268 | 1.00 (0.68–1.48) | 0.9900 |
| Season of booster vaccination |  |  |  |  |
| Mid-October to mid-April (Winter) | 1.00 |  | 1.00 |  |
| Mid-April to mid-October (Summer) | 0.63 (0.53–0.74) | <0.0001 | 0.55 (0.46–0.67) | <0.0001 |
| Previous infection |  |  |  |  |
| No evidence of previous infection | 1.00 |  | 1.00 |  |
| Previous infection before primary course of vaccination | 0.87 (0.64–1.17) | 0.3603 | 0.76 (0.56–1.04) | 0.0843 |
| Previous infection after primary course of vaccination | 0.30 (0.18–0.52) | <0.0001 | 0.26 (0.15–0.45) | <0.0001 |
| Probiotics |  |  |  |  |
| No | 1.00 |  | 1.00 |  |
| Yes | 0.77 (0.56–1.05) | 0.0948 | 0.81 (0.58–1.11) | 0.1853 |
| Fish oil, krill oil, or other omega-3 supplements |  |  |  |  |
| No | 1.00 |  | 1.00 |  |
| Yes | 0.79 (0.64–0.99) | 0.0409 | 0.83 (0.66–1.04) | 0.0983 |
| Cod liver oil supplements |  |  |  |  |
| No | 1.00 |  | 1.00 |  |
| Yes | 0.78 (0.60–1.02) | 0.0744 | 0.79 (0.61–1.04) | 0.0927 |
| Atopy |  |  |  |  |
| No | 1.00 |  | 1.00 |  |
| Yes | 0.85 (0.74–0.98) | 0.0258 | 0.90 (0.78–1.05) | 0.1776 |
| Diabetes |  |  |  |  |
| No diabetes | 1.00 |  | 1.00 |  |
| Pre-diabetes | 0.74 (0.48–1.12) | 0.1563 | 0.87 (0.57–1.33) | 0.5209 |
| Type 1 diabetes | 0.96 (0.48–1.93) | 0.9085 | 0.89 (0.44–1.79) | 0.7361 |
| Type 2 diabetes | 0.68 (0.47–0.98) | 0.0390 | 0.91 (0.53–1.56) | 0.7307 |
| Beta blockers |  |  |  |  |
| No | 1.00 |  | 1.00 |  |
| Yes | 0.74 (0.56–0.98) | 0.0342 | 0.86 (0.65–1.15) | 0.3078 |
| ACE inhibitors |  |  |  |  |
| No | 1.00 |  | 1.00 |  |
| Yes | 0.82 (0.65–1.03) | 0.0915 | 1.01 (0.79–1.29) | 0.9538 |
| Angiotensin receptor blockers |  |  |  |  |
| No | 1.00 |  | 1.00 |  |
| Yes | 0.76 (0.57–1.03) | 0.0745 | 0.91 (0.67–1.23) | 0.5437 |
| Thiazides |  |  |  |  |
| No | 1.00 |  | 1.00 |  |
| Yes | 0.51 (0.32–0.83) | 0.0067 | 0.60 (0.36–0.99) | 0.0466 |
| Metformin |  |  |  |  |
| No | 1.00 |  | 1.00 |  |
| Yes | 0.62 (0.39–0.99) | 0.0454 | 0.83 (0.42–1.63) | 0.5820 |

Risk factors associated with breakthrough infection at the 10% significance level in minimally adjusted models (ie, adjusted for age and sex) are shown. The fully adjusted analysis included all shown risk factors and was done in 10,590 participants, with 997 breakthrough infections. *p* values for quadratic, cubic, and quartic trends (where applicable) are shown in the appendix (table S3). ACE = angiotensin-converting-enzyme. ChAdOx1 = ChAdOx1 nCoV-19. HR = hazard ratio.

Examination of correlation matrices showed little evidence of collinearity between variables included in the fully adjusted models (data not shown). In the fully adjusted models, there was evidence of violation of the proportional hazards assumption for vaping (p=0.049) in the post-primary model and for vigorous physical activity (p=0.044), probiotics (p=0.046), and angiotensin receptor blockers (p=0.024) in the post-booster model. Stratifying by these variables did not substantially affect our estimates (appendix tables S4, S5).

## Discussion

In this large, prospective, national observational study, we found that lower levels of education, sharing a home with schoolchildren, and frequent visits to indoor public places other than shops were associated with increased risk of breakthrough SARS-CoV-2 infection, both after the primary course of vaccination and after receiving a booster dose. Older age and a previous SARS-CoV-2 infection were consistently associated with lower risk of breakthrough infection. We found higher risk of breakthrough infection after a ChAdOx1 primary course compared with a BNT162b2 primary course; this increased risk remained among ChAdOx1 recipients after a booster vaccination with mRNA-1273, when compared with participants who received BNT162b2 for both primary and booster doses.

Our study shows how the risk factors for SARS-CoV-2 infection after primary or booster vaccination can differ to those in unvaccinated populations. In pre-vaccination studies, high body-mass index (BMI),^22^ Asian ethnicity,^23^ and working in a healthcare setting^24^ have all been found to be risk factors for SARS-CoV-2 infection and COVID-19, including in the same COVIDENCE UK cohort studied here.^18^ By contrast, we observed no association with BMI, either after primary or booster vaccinations, and a reduced risk of breakthrough infection among healthcare workers after their primary vaccination. While underpowered to investigate ethnicity, previous studies on our cohort have consistently shown increased risk of pre-vaccination infection among south Asian participants,^18,19^ which has not remained after vaccination. Indeed, after a booster vaccination, we observed lower risk of breakthrough infection for participants of south Asian or mixed ethnicity compared with White participants after adjusting for previous infection. This supports both pre-vaccination and post-vaccination serology findings from the same cohort, showing higher antibody titres from natural infection and from vaccination in these ethnic groups than in White participants, independently of pre-vaccination serostatus^21^ and disease severity.^19^ Few studies to date have observed associations between ethnicity and breakthrough SARS-CoV-2 infection,^14^ although lack of diversity in vaccine clinical trials^25^ and missing data in some of the larger observational studies^2,9^ have hindered investigation. More large-scale studies with a more detailed breakdown of ethnicity are needed to understand the effects of booster vaccinations in ethnic subgroups.

Our findings highlight the important role that lifestyle and behaviours play in SARS-CoV-2 transmission after vaccination. Exposure to other people, both at home and in indoor public places, remained a strong predictor of breakthrough infection for fully vaccinated and boosted participants after adjusting for weekly SARS-CoV-2 incidence. In particular, sharing a home with schoolchildren— who represented a largely unvaccinated population for much of the study period—was consistently associated with a 40% increase in risk of infection. Notably, frequency of public transport use and visits to shops did not predict breakthrough infection in either model, suggesting that transmission largely occurs in settings of prolonged exposure, or where people are less likely to use masks, such as restaurants or bars. While we used pre-vaccination values for behaviours such as alcohol consumption and vaping to capture potential effects on immunogenicity,^26^ these values are unlikely to have changed greatly over follow-up and so may also reflect post-vaccination SARS-CoV-2 exposures, such as sharing of e-cigarettes.^27^

By contrast with studies on severe COVID-19 after vaccination,^16,17^ we found no indication of increased risk of breakthrough infection among people with comorbidities or among older participants, instead observing an inverse relationship with age. This finding is supported by other UK studies^2,28^ and matches the age patterns in infection rates observed over recent months in the UK.^29^ The fact that age remained a strong predictor in our cohort despite adjusting for behaviours such as public transport journeys and visits to indoor public places highlights the extent to which older adults, who are more likely to experience severe outcomes if infected,^30^ may still be reducing their social contacts compared with their younger counterparts.^31^ Similarly, the weak association seen between immunodeficiency and reduced risk of breakthrough infection in the post-primary cohort is likely to reflect more risk-averse behaviour in this group. These results suggest that despite adjusting for behaviours, we have not been able to capture them fully, and thus unmeasured or residual confounding may be operating in our study.

In our post-primary cohort, vaccine type was a strong predictor of breakthrough infection, with the weaker performance of ChAdOx1 mirroring studies on vaccine effectiveness in the general population^28^ and on post-vaccination serology.^21^ However, this difference seems to have been eradicated in our cohort after booster vaccination with BNT162b2. This may reflect both increased immunogenicity after boosting with BNT162b2, regardless of the primary vaccination course,^32^ and reduced effectiveness of BNT162b2 against the omicron variant,^5^ which became dominant in the UK in mid-December, 2021. Our results suggest that an mRNA-1273 booster might be less effective among ChAdOx1 recipients than a BNT162b2 booster, in contrast with studies that have found increased or similar immunogenicity in ChAdOx1-primed individuals receiving an mRNA-1273 booster versus a BNT162b2 booster.^32^ While our booster cohort contained substantially fewer mRNA-1273 recipients than BNT162b2 recipients, our large sample size lends strength to this finding, which should be investigated in future studies with longer follow-up.

We investigated several factors known to affect immune response to vaccination, such as season of vaccination,^33^ the inter-dose vaccine interval,^34^ and behaviours around the time of vaccination.^26^ We observed strong but inconsistent results for season of vaccination, with a higher risk of breakthrough infection among participants who received their first vaccine dose in the summer months (mid-April to mid-October) but lower risk among participants who received their booster dose in the summer months. Notably, the vast majority of first vaccinations in our cohort (14,353 [94.7%]) took place in the winter months, which is likely to have affected our results. It is likely that our season variable is additionally capturing time-varying factors not covered by our incidence and behavioural variables, such as more nuanced changes in behaviour owing to periods of restrictions and public health messaging, as well as changes in transmission risk arising from the emergence of new variants.

By contrast with studies showing that a longer inter-dose interval increases immunogenicity,^21,34,35^ we found a strong association between longer interval between first and second vaccine doses and increased risk of breakthrough infection. While a correlate of protection that translates immunogenicity findings into real-world protection has yet to be found, some preliminary studies suggest a longer interval is indeed protective.^36^ It is therefore possible that our finding of increased risk associated with longer inter-dose interval is the result of unmeasured confounding.

This study has several strengths. The size of the study, its population-based nature, and our inclusion of three of the most widely used vaccines for primary and booster vaccinations all increase the generalisability of our findings. The granular nature of the data used has allowed us to investigate a wide range of risk factors for breakthrough disease, while adjusting for weekly local SARS-CoV-2 incidence. Our use of monthly follow-up data means that we have been able to adjust for changing behaviours, rather than relying on baseline values that are unlikely to be representative of behaviours across different phases of the pandemic. Use of survey data has also allowed us to capture episodes of milder disease than those included in hospital-based studies, providing a clearer picture of post-vaccination transmission in the general population.

This study also has some limitations. First, we were unable to identify the strain of the infections, meaning that we could not dissect differences between the delta and omicron variants that were both dominant at different points in our study; however, more than 90% of the breakthrough infections in our post-booster analysis occurred after omicron became dominant in the UK, suggesting it largely captures the effects of that variant. Second, with very few hospitalisations, we did not have the power to assess risk factors for different levels of disease severity in vaccinated individuals. Third, as a self-selected cohort, several groups—including people younger than 30 years, people of lower socioeconomic status, and non-White ethnic groups—are under-represented in COVIDENCE UK. Lack of representativeness is not necessarily a barrier to valid scientific inference, however.^37^ Fourth, as with any observational study, it is possible that some of the associations we report can be explained by residual or unmeasured confounding; we have attempted to minimise this by adjusting for a comprehensive range of potential risk factors for infection, made possible by the large number of breakthrough infections recorded in our study (ie, >30 events per variable in each fully adjusted model). Finally, we explored several potential associations and cannot exclude the possibility that some achieved statistical significance as a result of type 1 error; however, by including the same predictors in post-primary and post-booster analyses, and presenting p-for-trend analyses for ordinal variables, we hope to have given readers the tools for a careful interpretation of our results.

In conclusion, primary and booster vaccinations have changed the landscape of the SARS-CoV-2 pandemic, attenuating the effects of risk factors for COVID-19 in unvaccinated populations, such as Asian ethnicity and BMI. We observed a clear difference between the efficacies of ChAdOx1 and BNT162b2 in fully vaccinated individuals, but the combination of BNT162b2 boosters and the omicron variant appears to have levelled the playing field. Key determinants of breakthrough infection remaining both after primary and booster vaccinations are therefore behaviours and lifestyles that affect individuals’ exposure to other people. As countries increasingly remove public health restrictions, we are truly entering an era of personal responsibility.

## Data Availability

De-identified participant data will be made available upon reasonable request to the corresponding author.

## Contributors

ARM wrote the study protocol, with input from HH, MT, and SOS. HH, MT, GAD, RAL, CJG, FK, AS, and ARM contributed to questionnaire development and design. HH co-ordinated and managed the study, with input from ARM, DAJ, MT, and SOS. HH, ARM, and SOS supported recruitment. GV, FT, MT, HH, and DAJ contributed to data management and coding medication data. GV and FT directly accessed and verified the data. Statistical analyses were done by GV, with input from ARM and FT. GV wrote the first draft of the report, with input from ARM. All authors read and approved the final manuscript. GV, DAJ, HH, FT, MT, SOS, and ARM had full access to all data in the study, and ARM had final responsibility for the decision to submit for publication.

## Declaration of interests

RAL has received grants from UKRI Medical Research Council, UKRI Economic and Social Research Council, Health Data Research UK, and Health and Care Research Wales. RAL is a member of the Welsh Government COVID-19 Technical Advisory group, in an unremunerated role. AS is a member of the Scottish Government’s Standing Committee on Pandemics, the Scottish Science Advisory Council, the UK Government’s New and Emerging Respiratory Virus Threats Risk Stratification Subgroup and the Department of Health and Social Care’s COVID-19 Therapeutics Modelling Group. He was a member of the Scottish Government Chief Medical Officer’s COVID-19 Advisory Group and AstraZeneca’s Thrombotic Thrombocytopenic Taskforce. All of AS’ roles are unremunerated. ARM declares receipt of funding to support vitamin D research from the following companies who manufacture or sell vitamin D supplements: Pharma Nord DSM Nutritional Products, Thornton & Ross, and Hyphens Pharma. ARM also declares support for attending meetings from the following companies who manufacture or sell vitamin D supplements: Pharma Nord and Abiogen Pharma. ARM also declares participation on the Data and Safety Monitoring Board for the Chair, DSMB, VITALITY trial (Vitamin D for Adolescents with HIV to reduce musculoskeletal morbidity and immunopathology). ARM also declares unpaid work as a Programme Committee member for the Vitamin D Workshop. ARM also declares receipt of vitamin D capsules for clinical trial use from Pharma Nord, Synergy Biologics, and Cytoplan. All other authors declare no competing interests.

## Acknowledgments

This study was supported by a grant from Barts Charity to ARM and CJG (MGU0466). The work was carried out with the support of BREATHE - The Health Data Research Hub for Respiratory Health (MC_PC_19004) in partnership with SAIL Databank. BREATHE is funded through the UK Research and Innovation (UKRI) Industrial Strategy Challenge Fund and delivered through Health Data Research UK. MT was supported by a grant from the Rosetrees Trust and The Bloom Foundation (M771) until May 2021 and has been supported by the Barts Charity since then (MGU0570). DAJ is supported by a Barts Charity Lectureship (ref MGU045). The views expressed are those of the authors and not necessarily those of Barts Charity, BREATHE, or Health Data Research UK. We thank all participants of COVIDENCE UK, and the following organisations who supported study recruitment: Asthma UK/British Lung Foundation, the British Heart Foundation, the British Lung Foundation, the British Obesity Society, Cancer Research UK, Diabetes UK, Future Publishing, Kidney Care UK, Kidney Wales, Mumsnet, the National Kidney Federation, the National Rheumatoid Arthritis Society, the North West London Health Research Register (DISCOVER), Primary Immunodeficiency UK, the Race Equality Foundation, SWM Health, the Terence Higgins Trust, and Vasculitis UK.

## Notes

### Funding Statement

Barts Charity, Health Data Research UK.

### Author Declarations

COVIDENCE UK was approved by Leicester South Research Ethics Committee (ref 20/EM/0117).

## References

1. Our World in Data. Coronavirus (COVID-19) vaccinations. 2022. https://ourworldindata.org/covid-vaccinations (accessed March 1, 2022).

2. Antonelli M, Penfold RS, Merino J, et al. Risk factors and disease profile of post-vaccination SARS-CoV-2 infection in UK users of the COVID Symptom Study app: a prospective, community-based, nested, case-control study. Lancet Infect Dis 2022; 22(1): 43–55.

3. Singanayagam A, Hakki S, Dunning J, et al. Community transmission and viral load kinetics of the SARS-CoV-2 delta (B.1.617.2) variant in vaccinated and unvaccinated individuals in the UK: a prospective, longitudinal, cohort study. Lancet Infect Dis 2022; 22: 183–95.

4. Feikin DR, Higdon MM, Abu-Raddad LJ, et al. Duration of effectiveness of vaccines against SARS-CoV-2 infection and COVID-19 disease: results of a systematic review and meta-regression. Lancet 2022; 399: 924–44.

5. Andrews N, Stowe J, Kirsebom F, et al. Effectiveness of COVID-19 vaccines against the Omicron (B.1.1.529) variant of concern. medRxiv 2021; published online Dec 14. DOI:2021.12.14.21267615 (preprint).

6. UK Health Security Agency. JCVI advice on COVID-19 booster vaccines for those aged 18 to 39 and a second dose for ages 12 to 15. 2021. https://www.gov.uk/government/news/jcvi-advice-on-covid-19-booster-vaccines-for-those-aged-18-to-39-and-a-second-dose-for-ages-12-to-15 (accessed March 1, 2022).

7. UK Government. Coronavirus (COVID-19) in the UK. 2022. https://coronavirus.data.gov.uk/ (accessed March 1, 2022).

8. Mackos D, Pruchnicka A. Factbox: Countries weigh need for COVID-19 booster shots. Reuters. Feb 21, 2022. https://www.reuters.com/business/healthcare-pharmaceuticals/countries-weigh-need-booster-covid-19-shots-2021-09-24 (accessed March 2, 2022).

9. Menni C, Klaser K, May A, et al. Vaccine side-effects and SARS-CoV-2 infection after vaccination in users of the COVID Symptom Study app in the UK: a prospective observational study. Lancet Infect Dis 2021; 21: 939–49.

10. Parameswaran A, Apsingi S, Eachempati KK, et al. Incidence and severity of COVID-19 infection post-vaccination: a survey among Indian doctors. Infection 2022; published online Feb 7. DOI:10.1007/s15010-022-01758-2.

11. Butt AA, Khan T, Yan P, Shaikh OS, Omer SB, Mayr F. Rate and risk factors for breakthrough SARS-CoV-2 infection after vaccination. J Infect 2021; 83: 237–79.

12. Sun J, Zheng Q, Madhira V, et al. Association Between Immune Dysfunction and COVID-19 Breakthrough Infection After SARS-CoV-2 Vaccination in the US. JAMA Int Med 2022; 182: 153–62.

13. Alishaq M, Nafady-Hego H, Jeremijenko A, et al. Risk factors for breakthrough SARS-CoV-2 infection in vaccinated healthcare workers. PLoS One 2021; 16: e0258820.

14. Liu C, Lee J, Ta C, et al. A Retrospective Analysis of COVID-19 mRNA Vaccine Breakthrough Infections - Risk Factors and Vaccine Effectiveness. medRxiv 2021; published online Oct 7. DOI:2021.10.05.21264583 (preprint).

15. Hollinghurst J, North L, Perry M, et al. COVID-19 infection risk amongst 14,104 vaccinated care home residents: a national observational longitudinal cohort study in Wales, UK, December 2020-March 2021. Age Ageing 2022; 51: afab223.

16. Hippisley-Cox J, Coupland CA, Mehta N, et al. Risk prediction of covid-19 related death and hospital admission in adults after covid-19 vaccination: national prospective cohort study. BMJ 2021; 374: n2244.

17. Grange Z, Buelo A, Sullivan C, et al. Characteristics and risk of COVID-19-related death in fully vaccinated people in Scotland. Lancet 2021; 398: 1799–800.

18. Holt H, Talaei M, Greenig M, et al. Risk factors for developing COVID-19: a population-based longitudinal study (COVIDENCE UK). Thorax 2021; published online Nov 30. DOI:10.1136/thoraxjnl-2021-217487.

19. Talaei M, Faustini S, Holt H, et al. Determinants of pre-vaccination antibody responses to SARS-CoV-2: a population-based longitudinal study (COVIDENCE UK). BMC Med 2022; 20: 87.

20. Department of Health and Social Care UG. Third primary COVID-19 vaccine dose for people who are immunosuppressed: JCVI advice. 2021.]https://www.gov.uk/government/publications/third-primary-covid-19-vaccine-dose-for-people-who-are-immunosuppressed-jcvi-advice (accessed March 9, 2022).

21. Jolliffe DA, Faustini SE, Holt H, et al. Determinants of antibody responses to two doses of ChAdOx1 nCoV-19 or BNT162b2 and a subsequent booster dose of BNT162b2 or mRNA-1273: population-based cohort study (COVIDENCE UK). medRxiv 2022; published online Feb 15. DOI:2022.02.14.22270930 (preprint).

22. de Lusignan S, Dorward J, Correa A, et al. Risk factors for SARS-CoV-2 among patients in the Oxford Royal College of General Practitioners Research and Surveillance Centre primary care network: a cross-sectional study. Lancet Infect Dis 2020; 20: 1034–42.

23. Ward H, Atchison C, Whitaker M, et al. SARS-CoV-2 antibody prevalence in England following the first peak of the pandemic. Nat Commun 2021; 12: 905.

24. Chadeau-Hyam M, Bodinier B, Elliott J, et al. Risk factors for positive and negative COVID-19 tests: a cautious and in-depth analysis of UK biobank data. Int J Epidemiol 2020; 49: 1454–67.

25. Jethwa H, Wong R, Abraham S. Covid-19 vaccine trials: Ethnic diversity and immunogenicity. Vaccine 2021; 39: 3541–43.

26. Zimmermann P, Curtis N. Factors That Influence the Immune Response to Vaccination. Clin Microbiol Rev 2019; 32: e00084–18.

27. Besaratinia A, Tommasi S. The consequential impact of JUUL on youth vaping and the landscape of tobacco products: The state of play in the COVID-19 era. Preventive Medicine Reports 2021; 22: 101374.

28. Chadeau-Hyam M, Wang H, Eales O, et al. SARS-CoV-2 infection and vaccine effectiveness in England (REACT-1): a series of cross-sectional random community surveys. Lancet Respir Med 2022; published online Jan 24. DOI:10.1016/S2213-2600(21)00542-7.

29. Office for National Statistics. Coronavirus (COVID-19) Infection Survey, UK: 25 February 2022. 2022. https://www.ons.gov.uk/peoplepopulationandcommunity/healthandsocialcare/conditionsanddiseases/bulletins/coronaviruscovid19infectionsurveypilot/25february2022 (accessed Feb 25, 2022).

30. Williamson EJ, Walker AJ, Bhaskaran K, et al. Factors associated with COVID-19-related death using OpenSAFELY. Nature 2020; 584: 430–36.

31. Jarvis CI, Gimma A, Wong KL, et al. Social contacts in the UK from the CoMix social contact survey Report for survey week 101 - Final report. March 8, 2022. https://cmmid.github.io/topics/covid19/comix-reports.html (accessed March 9, 2022).

32. Munro APS, Janani L, Cornelius V, et al. Safety and immunogenicity of seven COVID-19 vaccines as a third dose (booster) following two doses of ChAdOx1 nCov-19 or BNT162b2 in the UK (COV-BOOST): a blinded, multicentre, randomised, controlled, phase 2 trial. Lancet 2021; 398: 2258–76.

33. Dopico XC, Evangelou M, Ferreira RC, et al. Widespread seasonal gene expression reveals annual differences in human immunity and physiology. Nat Commun 2015; 6: 7000.

34. Payne RP, Longet S, Austin JA, et al. Immunogenicity of standard and extended dosing intervals of BNT162b2 mRNA vaccine. Cell 2021; 184: 5699–714.e11.

35. Flaxman A, Marchevsky NG, Jenkin D, et al. Reactogenicity and immunogenicity after a late second dose or a third dose of ChAdOx1 nCoV-19 in the UK: a substudy of two randomised controlled trials (COV001 and COV002). Lancet 2021; 398: 981–90.

36. Fisman DN, Lee N, Tuite AR. Timing of Breakthrough Infection Risk After Vaccination Against SARS-CoV-2. medRxiv 2022; published online Jan 5. DOI:2022.01.04.22268773 (preprint).

37. Rothman KJ, Gallacher JEJ, Hatch EE. Why representativeness should be avoided. Int J Epidemiol 2013; 42: 1012–14.

