## Appendix for "Risk factors for SARS-CoV-2 infection after primary vaccination with ChAdOx1 nCoV-19 or BNT1262b2 and after booster vaccination with BNT1262b2 or mRNA-1273: a population-based cohort study (COVIDENCE UK)"

### ***Table S1:* Factors not associated with breakthrough infection in the post-primary cohort, after adjustment for age and sex**

|  |  | | **HR (95% CI)** | | | **p value** |
| --- | --- | --- | --- | --- | --- | --- |
|  | Ethnicity | |  | | |  |
|  | White | | 1.00 | | |  |
|  | Mixed, multiple, or other ethnic groups | | 1.01 (0.72–1.41) | | | 0.9742 |
|  | South Asian | | 0.75 (0.47–1.18) | | | 0.2099 |
|  | Black, African, Caribbean, or Black British | | 0.42 (0.14–1.31) | | | 0.1340 |
|  | Quartiles of IMD rank | |  | | |  |
|  | Q4 (least deprived) | | 1.00 | | |  |
|  | Q3 | | 0.89 (0.75–1.06) | | | 0.1903 |
|  | Q2 | | 1.05 (0.89–1.25) | | | 0.5369 |
|  | Q1 (most deprived) | | 0.90 (0.76–1.07) | | | 0.2333 |
|  | Claiming Universal Credit | |  | | |  |
|  | No | | 1.00 | | |  |
|  | Yes | | 1.10 (0.78–1.54) | | | 0.5883 |
|  | Dog at home | |  | | |  |
|  | No | | 1.00 | | |  |
|  | Yes | | 0.94 (0.81–1.08) | | | 0.3802 |
|  | Smoking status | |  | | |  |
|  | No | | 1.00 | | |  |
|  | Yes | | 1.12 (0.87–1.45) | | | 0.3863 |
|  | Environmental tobacco smoke exposure | |  | | |  |
|  | No | | 1.00 | | |  |
|  | Yes | | 1.03 (0.68–1.58) | | | 0.8813 |
|  | Travel outside of the UK since last questionnaire | |  | | |  |
|  | No | | 1.00 | | |  |
|  | Yes | | 1.07 (0.91–1.25) | | | 0.4161 |
|  | Weekly journeys on public transport | |  | | |  |
|  | 0 | | 1.00 | | |  |
|  | 1–5 | | 1.01 (0.87–1.17) | | | 0.9186 |
|  | ≥6 | | 1.01 (0.81–1.26) | | | 0.9126 |
|  | Lower impact physical exercise, h per week | |  | | |  |
|  | 0 | | 0.97 (0.84–1.13) | | | 0.7236 |
|  | 1 | | 0.92 (0.76–1.12) | | | 0.4224 |
|  | ≥2 | | 1.00 | | |  |
|  | Light physical exercise, h per week | |  | | |  |
|  | 0–2 | | 1.01 (0.84–1.21) | | | 0.8984 |
|  | 3–5 | | 1.13 (0.95–1.34) | | | 0.1568 |
|  | 7–9 | | 1.06 (0.88–1.28) | | | 0.5331 |
|  | ≥10 | | 1.00 | | |  |
|  | Vigorous physical exercise,  h per week | |  | | |  |
|  | 0 | | 0.95 (0.80–1.13) | | | 0.5615 |
|  | 1–3 | | 0.99 (0.84–1.18) | | | 0.9235 |
|  | ≥4 | | 1.00 | | |  |
|  | Actual sleep, h/night | |  | | |  |
|  | ≤5 | | 0.96 (0.78–1.19) | | | 0.7239 |
|  | 6 | | 1.09 (0.94–1.27) | | | 0.2590 |
|  | 7 | | 1.00 | | |  |
|  | ≥8 | | 0.91 (0.77–1.06) | | | 0.2208 |
|  | Multivitamin supplements | |  | | |  |
|  | No | | 1.00 | | |  |
|  | Yes | | 1.08 (0.94–1.24) | | | 0.2866 |
|  | Vitamin A supplements | |  | | |  |
|  | No | | 1.00 | | |  |
|  | Yes | | 0.92 (0.48–1.77) | | | 0.7979 |
|  | Vitamin C supplements | |  | | |  |
|  | No | | 1.00 | | |  |
|  | Yes | | 1.06 (0.88–1.28) | | | 0.5165 |
|  |  | | **HR (95% CI)** | | | **p value** |
|  | Vitamin D supplements | |  | | |  |
|  | No | | 1.00 | | |  |
|  | Yes | | 1.02 (0.90–1.15) | | | 0.7521 |
|  | Zinc supplements | |  | | |  |
|  | No | | 1.00 | | |  |
|  | Yes | | 1.05 (0.83–1.32) | | | 0.6946 |
|  | Selenium supplements | |  | | |  |
|  | No | | 1.00 | | |  |
|  | Yes | | 1.07 (0.63–1.82) | | | 0.7900 |
|  | Iron supplements | |  | | |  |
|  | No | | 1.00 | | |  |
|  | Yes | | 0.77 (0.55–1.09) | | | 0.1379 |
|  | Probiotics | |  | | |  |
|  | No | | 1.00 | | |  |
|  | Yes | | 0.97 (0.75–1.25) | | | 0.7923 |
|  | Fish oil, krill oil, or other omega-3 supplements | |  | | |  |
|  | No | | 1.00 | | |  |
|  | Yes | | 0.98 (0.80–1.19) | | | 0.8024 |
|  | Cod liver oil supplements | |  | | |  |
|  | No | | 1.00 | | |  |
|  | Yes | | 1.02 (0.80–1.28) | | | 0.8992 |
|  | Garlic or allicin supplements | |  | | |  |
|  | No | | 1.00 | | |  |
|  | Yes | | 0.88 (0.51–1.51) | | | 0.6336 |
|  | Daily portions of dairy products or calcium-fortified alternatives | |  | | |  |
|  | 0 | | 1.00 | | |  |
|  | 1 | | 0.95 (0.68–1.32) | | | 0.7583 |
|  | 2–3 | | 1.01 (0.73–1.38) | | | 0.9610 |
|  | ≥4 | | 1.00 (0.72–1.38) | | | 0.9932 |
|  | Food choice | |  | | |  |
|  | None | | 1.00 | | |  |
|  | Vegetarian | | 0.98 (0.74–1.29) | | | 0.8779 |
|  | Vegan | | 1.16 (0.75–1.79) | | | 0.4991 |
|  | BMI, kg/m² | |  | | |  |
|  | <25 | | 1.00 | | |  |
|  | 25 to <30 | | 1.02 (0.88–1.17) | | | 0.8154 |
|  | ≥30 | | 1.11 (0.95–1.30) | | | 0.1742 |
|  | Heart disease | |  | | |  |
|  | No | | 1.00 | | |  |
|  | Yes | | 0.96 (0.66–1.40) | | | 0.8513 |
|  | Arterial disease | |  | | |  |
|  | No | | 1.00 | | |  |
|  | Yes | | 0.92 (0.66–1.28) | | | 0.6178 |
|  | Periodontitis | |  | | |  |
|  | No | | 1.00 | | |  |
|  | Yes | | 1.03 (0.90–1.19) | | | 0.6583 |
|  | Major neurological conditions | |  | | |  |
|  | No | | 1.00 | | |  |
|  | Yes | | 1.10 (0.74–1.62) | | | 0.6484 |
|  | Asthma | |  | | |  |
|  | No | | 1.00 | | |  |
|  | Yes | | 1.02 (0.87–1.19) | | | 0.8178 |
|  | COPD | |  | | |  |
|  | No | | 1.00 | | |  |
|  | Yes | | 0.74 (0.42–1.27) | | | 0.2723 |
|  | Atopy | |  | | |  |
|  | No | | 1.00 | | |  |
|  | Yes | | 0.96 (0.84–1.10) | | | 0.5456 |
|  | Cancer | |  | | |  |
|  | Never | | 1.00 | | |  |
|  | Past (cured or in remission) | | 0.92 (0.72–1.18) | | | 0.5302 |
|  | Present (active treatment) | | 1.10 (0.55–2.22) | | | 0.7808 |
|  | |  | | **HR (95% CI)** | **p value** | |
|  | Diabetes types | |  | | |  |
|  | No diabetes | | 1.00 | | |  |
|  | Pre-diabetes | | 0.94 (0.63–1.40) | | | 0.7481 |
|  | Type 1 diabetes | | 0.74 (0.33–1.66) | | | 0.4656 |
|  | Type 2 diabetes | | 0.96 (0.69–1.34) | | | 0.8088 |
|  | | Beta-2 adrenergic agonists | |  |  | |
|  | | No | | 1.00 |  | |
|  | | Yes | | 0.87 (0.71–1.07) | 0.1925 | |
|  | | Beta blockers | |  |  | |
|  | | No | | 1.00 |  | |
|  | | Yes | | 1.13 (0.88–1.45) | 0.3541 | |
|  | | Statins | |  |  | |
|  | | No | | 1.00 |  | |
|  | | Yes | | 0.98 (0.80–1.19) | 0.8140 | |
|  | | ACE inhibitors | |  |  | |
|  | | No | | 1.00 |  | |
|  | | Yes | | 0.84 (0.66–1.07) | 0.1593 | |
|  | | Proton pump inhibitors | |  |  | |
|  | | No | | 1.00 |  | |
|  | | Yes | | 0.98 (0.81–1.18) | 0.8192 | |
|  | | H2-receptor antagonists | |  |  | |
|  | | No | | 1.00 |  | |
|  | | Yes | | 0.77 (0.32–1.84) | 0.5512 | |
|  | | Inhaled corticosteroids | |  |  | |
|  | | No | | 1.00 |  | |
|  | | Yes | | 0.89 (0.69–1.15) | 0.3720 | |
|  | | Bronchodilators | |  |  | |
|  | | No | | 1.00 |  | |
|  | | Yes | | 0.88 (0.72–1.08) | 0.2312 | |
|  | | Systemic immunosuppressants | |  |  | |
|  | | No | | 1.00 |  | |
|  | | Yes | | 0.99 (0.74–1.31) | 0.9215 | |
|  | | Angiotensin receptor blockers | |  |  | |
|  | | No | | 1.00 |  | |
|  | | Yes | | 1.03 (0.77–1.36) | 0.8606 | |
|  | | SSRIs | |  |  | |
|  | | No | | 1.00 |  | |
|  | | Yes | | 0.96 (0.76–1.20) | 0.6929 | |
|  | | Non-SSRI antidepressants | |  |  | |
|  | | No | | 1.00 |  | |
|  | | Yes | | 0.83 (0.60–1.15) | 0.2688 | |
|  | | Thiazides | |  |  | |
|  | | No | | 1.00 |  | |
|  | | Yes | | 0.78 (0.50–1.22) | 0.2848 | |
|  | | Vitamin K antagonists | |  |  | |
|  | | No | | 1.00 |  | |
|  | | Yes | | 0.79 (0.33–1.91) | 0.6023 | |
|  | | SGLT2 inhibitors | |  |  | |
|  | | No | | 1.00 |  | |
|  | | Yes | | 1.70 (0.85–3.42) | 0.1351 | |
|  | | Metformin | |  |  | |
|  | | No | | 1.00 |  | |
|  | | Yes | | 1.17 (0.81–1.69) | 0.3976 | |
|  | | Bisphosphonates | |  |  | |
|  | | No | | 1.00 |  | |
|  | | Yes | | 0.70 (0.38–1.31) | 0.2681 | |
|  | | Anti-platelet drugs | |  |  | |
|  | | No | | 1.00 |  | |
|  | | Yes | | 1.05 (0.80–1.39) | 0.7064 | |
|  | | Sex hormone therapy | |  |  | |
|  | | No | | 1.00 |  | |
|  | | Yes | | 1.01 (0.81–1.26) | 0.9154 | |
|  | | Aspirin | |  |  | |
|  | | No | | 1.00 |  | |
|  | | Yes | | 1.14 (0.85–1.54) | 0.3843 | |
|  | |  | | **HR (95% CI)** | **p value** | |
|  | | Paracetamol | |  |  | |
|  | | No | | 1.00 |  | |
|  | | Yes | | 1.14 (0.85–1.52) | 0.3780 | |
|  | | BCG vaccinated | |  |  | |
|  | | No | | 1.00 |  | |
|  | | Yes | | 0.99 (0.83–1.19) | 0.9448 | |

ACE = angiotensin-converting-enzyme. BCG = Bacille Calmette Guérin. BMI = body-mass index. COPD = chronic obstructive pulmonary disease. HR = hazard ratio. IMD = Index of Multiple Deprivation. SLT2 = sodium-glucose co-transporter-2. SSRI = selective serotonin reuptake inhibitors.

### ***Table S2:* Factors not associated with breakthrough infection in the post-booster cohort, after adjustment for age and sex**

|  |  | **HR (95% CI)** | **p value** |
| --- | --- | --- | --- |
|  | Number of people per bedroom |  |  |
|  | <1 | 1.00 |  |
|  | 1 to <2 | 1.09 (0.94–1.26) | 0.2673 |
|  | ≥2 | 1.36 (0.89–2.10) | 0.1579 |
|  | Quartiles of IMD rank |  |  |
|  | Q4 (least deprived) | 1.00 |  |
|  | Q3 | 0.91 (0.77–1.08) | 0.3048 |
|  | Q2 | 1.00 (0.84–1.19) | 0.9882 |
|  | Q1 (most deprived) | 0.99 (0.83–1.18) | 0.8787 |
|  | Claiming Universal Credit |  |  |
|  | No | 1.00 |  |
|  | Yes | 0.94 (0.59–1.50) | 0.7891 |
|  | Dog at home |  |  |
|  | No | 1.00 |  |
|  | Yes | 1.08 (0.93–1.25) | 0.3134 |
|  | Smoking status |  |  |
|  | No | 1.00 |  |
|  | Yes | 0.93 (0.67–1.30) | 0.6638 |
|  | Vaping status |  |  |
|  | No | 1.00 |  |
|  | Yes | 1.10 (0.75–1.60) | 0.6271 |
|  | Environmental tobacco smoke exposure |  |  |
|  | No | 1.00 |  |
|  | Yes | 1.15 (0.73–1.81) | 0.5421 |
|  | Lower impact physical exercise, h per week |  |  |
|  | 0 | 1.05 (0.89–1.24) | 0.5468 |
|  | 1 | 1.13 (0.93–1.37) | 0.2314 |
|  | ≥2 | 1.00 |  |
|  | Light physical exercise,  h per week |  |  |
|  | 0–2 | 0.91 (0.76–1.10) | 0.3404 |
|  | 3–5 | 1.01 (0.86–1.18) | 0.9084 |
|  | 7–9 | 0.92 (0.77–1.09) | 0.3409 |
|  | ≥10 | 1.00 |  |
|  | Multivitamin supplements |  |  |
|  | No | 1.00 |  |
|  | Yes | 0.89 (0.76–1.04) | 0.1486 |
|  | Vitamin A supplements |  |  |
|  | No | 1.00 |  |
|  | Yes | 0.96 (0.43–2.13) | 0.9116 |
|  | Vitamin C supplements |  |  |
|  | No | 1.00 |  |
|  | Yes | 1.00 (0.81–1.24) | 0.9857 |
|  | Vitamin D supplements |  |  |
|  | No | 1.00 |  |
|  | Yes | 0.99 (0.88–1.12) | 0.9008 |
|  | Zinc supplements |  |  |
|  | No | 1.00 |  |
|  | Yes | 0.86 (0.65–1.13) | 0.2849 |
|  | Selenium supplements |  |  |
|  | No | 1.00 |  |
|  | Yes | 0.80 (0.40–1.61) | 0.5362 |
|  | Iron supplements |  |  |
|  | No | 1.00 |  |
|  | Yes | 0.82 (0.56–1.21) | 0.3167 |
|  | Garlic or allicin supplements |  |  |
|  | No | 1.00 |  |
|  | Yes | 0.87 (0.50–1.50) | 0.6163 |

|  | |  | | | **HR (95% CI)** | | **p value** |
| --- | --- | --- | --- | --- | --- | --- | --- |
|  | | Daily portions of fruit, vegetable, and salad | |  | | |  |
|  | | 0–2 | | 1.00 | | |  |
|  | | 3–4 | | 1.00 (0.82–1.22) | | | 0.9907 |
|  | | 5 | | 0.90 (0.72–1.13) | | | 0.3732 |
|  | | ≥6 | | 0.96 (0.78–1.18) | | | 0.7044 |
|  | | Daily portions of dairy products or calcium-fortified alternatives | | |  | |  |
|  | | 0 | | | 1.29 (0.92–1.80) | | 0.1395 |
|  | | 1 | | | 0.97 (0.82–1.16) | | 0.7647 |
|  | | 2–3 | | | 0.97 (0.83–1.12) | | 0.6730 |
|  | | ≥4 | | | 1.00 | |  |
|  | | Food choice | | |  | |  |
|  | | None | | | 1.00 | |  |
|  | | Vegetarian | | | 0.81 (0.59–1.12) | | 0.2017 |
|  | | Vegan | | | 0.79 (0.45–1.40) | | 0.4197 |
|  | | General health | | |  | |  |
|  | | Excellent | | | 1.00 | |  |
|  | | Very good | | | 1.04 (0.86–1.25) | | 0.7048 |
|  | | Good | | | 0.99 (0.81–1.21) | | 0.9235 |
|  | | Fair | | | 0.82 (0.64–1.05) | | 0.1138 |
|  | | Poor | | | 0.90 (0.61–1.33) | | 0.6075 |
|  | | BMI, kg/m² | | |  | |  |
|  | | <25 | | | 1.00 | |  |
|  | | 25 to <30 | | | 1.08 (0.93–1.24) | | 0.3118 |
|  | | ≥30 | | | 0.98 (0.82–1.16) | | 0.7769 |
|  | | Heart disease | | |  | |  |
|  | | No | | | 1.00 | |  |
|  | | Yes | | | 0.92 (0.65–1.32) | | 0.6648 |
|  | | Arterial disease | | |  | |  |
|  | | No | | | 1.00 | |  |
|  | | Yes | | | 0.89 (0.65–1.21) | | 0.4588 |
|  | | Periodontitis | | |  | |  |
|  | | No | | | 1.00 | |  |
|  | | Yes | | | 1.01 (0.88–1.16) | | 0.8682 |
|  | | Hypertension | | |  | |  |
|  | | No | | | 1.00 | |  |
|  | | Yes | | | 0.88 (0.75–1.04) | | 0.1390 |
|  | | Immunodeficiency | | |  | |  |
|  | | No | | | 1.00 | |  |
|  | | Yes | | | 1.22 (0.67–2.21) | | 0.5145 |
|  | | Major neurological conditions | | |  | |  |
|  | | No | | | 1.00 | |  |
|  | | Yes | | | 0.72 (0.46–1.13) | | 0.1570 |
|  | | Asthma | | |  | |  |
|  | | No | | | 1.00 | |  |
|  | | Yes | | | 0.99 (0.84–1.16) | | 0.8976 |
|  | | COPD | | |  | |  |
|  | | No | | | 1.00 | |  |
|  | | Yes | | | 0.89 (0.55–1.41) | | 0.6107 |
|  | | Cancer | | |  | |  |
|  | | Never | | | 1.00 | |  |
|  | | Past (cured or in remission) | | | 0.88 (0.69–1.10) | | 0.2636 |
|  | | Present (active treatment) | | | 0.88 (0.44–1.77) | | 0.7230 |
|  | | Beta-2 adrenergic agonists | | |  | |  |
|  | | No | | | 1.00 | |  |
|  | | Yes | | | 0.87 (0.70–1.08) | | 0.2063 |
|  | | Statins | | |  | |  |
|  | | No | | | 1.00 | |  |
|  | | Yes | | | 0.95 (0.79–1.13) | | 0.5465 |
|  | | Proton pump inhibitors | | |  | |  |
|  | | No | | | 1.00 | |  |
|  | | Yes | | | 0.86 (0.72–1.04) | | 0.1183 |
|  |  | | **HR (95% CI)** | | | **p value** | |
|  | H2-receptor antagonists | |  | | |  | |
|  | No | | 1.00 | | |  | |
|  | Yes | | 0.88 (0.40–1.97) | | | 0.7605 | |
|  | Inhaled corticosteroids | |  | | |  | |
|  | No | | 1.00 | | |  | |
|  | Yes | | 0.97 (0.76–1.24) | | | 0.8216 | |
|  | Bronchodilators | |  | | |  | |
|  | No | | 1.00 | | |  | |
|  | Yes | | 0.89 (0.72–1.10) | | | 0.2723 | |
|  | Systemic immunosuppressants | |  | | |  | |
|  | No | | 1.00 | | |  | |
|  | Yes | | 0.89 (0.68–1.17) | | | 0.4038 | |
|  | SSRIs | |  | | |  | |
|  | No | | 1.00 | | |  | |
|  | Yes | | 0.83 (0.63–1.08) | | | 0.1695 | |
|  | Non-SSRI antidepressants | |  | | |  | |
|  | No | | 1.00 | | |  | |
|  | Yes | | 0.89 (0.65–1.22) | | | 0.4770 | |
|  | Calcium channel blockers | |  | | |  | |
|  | No | | 1.00 | | |  | |
|  | Yes | | 0.96 (0.77–1.20) | | | 0.7485 | |
|  | Vitamin K antagonists | |  | | |  | |
|  | No | | 1.00 | | |  | |
|  | Yes | | 0.92 (0.43–1.93) | | | 0.8155 | |
|  | SGLT2 inhibitors | |  | | |  | |
|  | No | | 1.00 | | |  | |
|  | Yes | | 0.21 (0.03–1.47) | | | 0.1152 | |
|  | Anticholinergics | |  | | |  | |
|  | No | | 1.00 | | |  | |
|  | Yes | | 0.82 (0.61–1.09) | | | 0.1723 | |
|  | Bisphosphonates | |  | | |  | |
|  | No | | 1.00 | | |  | |
|  | Yes | | 0.68 (0.40–1.16) | | | 0.1564 | |
|  | Anti-platelet drugs | |  | | |  | |
|  | No | | 1.00 | | |  | |
|  | Yes | | 0.94 (0.72–1.22) | | | 0.6252 | |
|  | Sex hormone therapy | |  | | |  | |
|  | No | | 1.00 | | |  | |
|  | Yes | | 1.12 (0.90–1.39) | | | 0.2975 | |
|  | Aspirin | |  | | |  | |
|  | No | | 1.00 | | |  | |
|  | Yes | | 0.91 (0.68–1.23) | | | 0.5548 | |
|  | Paracetamol | |  | | |  | |
|  | No | | 1.00 | | |  | |
|  | Yes | | 0.83 (0.60–1.15) | | | 0.2619 | |
|  | BCG vaccinated | |  | | |  | |
|  | No | | 1.00 | | |  | |
|  | Yes | | 1.08 (0.89–1.32) | | | 0.4448 | |

ACE = angiotensin-converting-enzyme. BCG = Bacille Calmette Guérin. BMI = body-mass index. COPD = chronic obstructive pulmonary disease. HR = hazard ratio. IMD = Index of Multiple Deprivation. SLT2 = sodium-glucose co-transporter-2. SSRI = Selective serotonin reuptake inhibitors.

|  |  | **p for trend** | |
| --- | --- | --- | --- |
|  |  | Post-primary | Post-booster |
|  | Highest education level attained |  |  |
|  | Linear | <0.0001 | 0.0043 |
|  | Quadratic | 0.1653 | 0.6141 |
|  | Cubic | 0.3016 | 0.7931 |
|  | Number of people per bedroom |  |  |
|  | Linear | 0.0131 | .. |
|  | Quadratic | 0.2812 | .. |
|  | Multigenerational households |  |  |
|  | Linear | 0.3789 | 0.6556 |
|  | Quadratic | 0.7161 | 0.4656 |
|  | Alcohol consumption, units per week |  |  |
|  | Linear | 0.0460 | 0.0234 |
|  | Quadratic | 0.8199 | 0.2374 |
|  | Cubic | 0.6002 | 0.1653 |
|  | Weekly journeys on public transport |  |  |
|  | Linear | .. | 0.4084 |
|  | Quadratic | .. | 0.4075 |
|  | Weekly visits to shops |  |  |
|  | Linear | 0.5823 | 0.0418 |
|  | Quadratic | 0.3498 | 0.4605 |
|  | Cubic | 0.5353 | 0.5617 |
|  | Weekly visits to other indoor public places (not shops) |  |  |
|  | Linear | 0.0005 | 0.0021 |
|  | Quadratic | 0.9827 | 0.4505 |
|  | Vigorous physical exercise, h per week |  |  |
|  | Linear | .. | 0.2058 |
|  | Quadratic | .. | 0.6240 |
|  | Actual sleep, h/night |  |  |
|  | Linear | .. | 0.0547 |
|  | Quadratic | .. | 0.0595 |
|  | Cubic | .. | 0.1601 |
|  | Daily portions of fruit, vegetables, and salad |  |  |
|  | Linear | 0.1651 | .. |
|  | Quadratic | 0.7277 | .. |
|  | Cubic | 0.2591 | .. |
|  | General health |  |  |
|  | Linear | 0.2918 | .. |
|  | Quadratic | 0.2276 | .. |
|  | Cubic | 0.0352 | .. |
|  | Quartic | 0.9671 | .. |

### ***Table S3:* Trend analysis**

### ***Table S4:* Risk factors for breakthrough infection in the pre-booster cohort, stratified by vaping status**

|  |  | **HR (95% CI)** | **p value** |
| --- | --- | --- | --- |
|  | Age, years | 0.96 (0.96–0.97) | <0.0001 |
|  | Sex |  |  |
|  | Female | 1.00 |  |
|  | Male | 1.03 (0.89–1.20) | 0.6514 |
|  | Highest educational level attained |  |  |
|  | Post-grad | 1.00 |  |
|  | College or university | 1.10 (0.95–1.28) | 0.1925 |
|  | Higher or further (A levels) | 1.18 (0.96–1.44) | 0.1161 |
|  | Primary or secondary | 1.67 (1.35–2.07) | <0.0001 |
|  | Frontline worker |  |  |
|  | No | 1.00 |  |
|  | Non-health | 1.19 (1.00–1.42) | 0.0512 |
|  | Health | 0.72 (0.56–0.92) | 0.0094 |
|  | Housing |  |  |
|  | Owns own home | 1.00 |  |
|  | Mortgage | 1.11 (0.94–1.31) | 0.2331 |
|  | Privately renting | 0.83 (0.63–1.09) | 0.1843 |
|  | Renting from council | 1.27 (0.90–1.79) | 0.1696 |
|  | Other | 0.86 (0.61–1.21) | 0.3802 |
|  | Number of people per bedroom |  |  |
|  | <1 | 1.00 |  |
|  | 1 to <2 | 1.16 (1.00–1.35) | 0.0550 |
|  | ≥2 | 1.66 (1.19–2.31) | 0.0029 |
|  | Multigenerational households |  |  |
|  | Living alone | 0.91 (0.73–1.15) | 0.4368 |
|  | Single generation | 1.00 |  |
|  | Two or more generations | 1.03 (0.85–1.24) | 0.7916 |
|  | Shares home with schoolchildren (5–15 years) |  |  |
|  | No | 1.00 |  |
|  | Yes | 1.41 (1.12–1.77) | 0.0031 |
|  | Shares home with working-age adult (16–64 year) |  |  |
|  | No | 1.00 |  |
|  | Yes | 1.02 (0.85–1.23) | 0.7961 |
|  | Alcohol consumption, units per week |  |  |
|  | 0 | 1.00 |  |
|  | 1–7 | 1.05 (0.90–1.23) | 0.5247 |
|  | 8–14 | 1.17 (0.98–1.40) | 0.0887 |
|  | ≥15 | 1.18 (0.97–1.43) | 0.1066 |
|  | Any visits to or from other households in past week |  |  |
|  | No | 1.00 |  |
|  | Yes | 1.18 (1.00–1.38) | 0.0467 |
|  | Weekly visits to shops |  |  |
|  | 0 | 1.00 |  |
|  | 1 | 1.04 (0.75–1.46) | 0.7998 |
|  | 2–3 | 1.15 (0.85–1.56) | 0.3557 |
|  | ≥4 | 1.10 (0.81–1.49) | 0.5440 |
|  | Weekly visits to other indoor public places (not shops) |  |  |
|  | 0 | 1.00 |  |
|  | 1–2 | 1.18 (0.99–1.40) | 0.0650 |
|  | ≥3 | 1.38 (1.15–1.67) | 0.0005 |
|  | Weekly SARS-CoV-2 incidence (per 1000 people) | 1.05 (1.03–1.06) | <0.0001 |
|  | Inter-vaccine interval, weeks | 1.08 (1.07–1.10) | <0.0001 |
|  | Primary vaccination course |  |  |
|  | ChAdOx1 | 1.62 (1.40–1.87) | <0.0001 |
|  | BNT162b2 | 1.00 |  |
|  | Season of first vaccination |  |  |
|  | Mid-October to mid-April (Winter) | 1.00 |  |
|  | Mid-April to mid-October (Summer) | 2.15 (1.72–2.69) | <0.0001 |
|  | Previous infection |  |  |
|  | No | 1.00 |  |
|  | Yes | 0.55 (0.40–0.75) | 0.0002 |
|  | Quartiles of daily fruit, vegetable, and salad intake |  |  |
|  | 0–2 | 1.00 |  |
|  | 3–4 | 1.15 (0.94–1.40) | 0.1831 |
|  | 5 | 1.09 (0.88–1.37) | 0.4280 |
|  | ≥6 | 1.20 (0.98–1.47) | 0.0834 |
|  | General health |  |  |
|  | Excellent | 1.00 |  |
|  | Very good | 0.86 (0.73–1.02) | 0.0908 |
|  | Good | 0.97 (0.80–1.16) | 0.7225 |
|  | Fair | 1.09 (0.86–1.37) | 0.4755 |
|  | Poor | 0.93 (0.64–1.36) | 0.7157 |
|  | Asthma |  |  |
|  | No | 1.00 |  |
|  | Yes | 1.14 (0.97–1.35) | 0.1225 |
|  | Immunodeficiency |  |  |
|  | No | 1.00 |  |
|  | Yes | 0.17 (0.02–1.23) | 0.0800 |
|  | Calcium channel blockers |  |  |
|  | No | 1.00 |  |
|  | Yes | 0.85 (0.65–1.10) | 0.2082 |
|  | Anticholinergics |  |  |
|  | No | 1.00 |  |
|  | Yes | 0.66 (0.47–0.94) | 0.0215 |

ACE = angiotensin-converting-enzyme. BCG = Bacille Calmette Guérin. BMI = body-mass index. COPD = chronic obstructive pulmonary disease. HR = hazard ratio. IMD = Index of Multiple Deprivation. SLT2 = sodium-glucose co-transporter-2. SSRI = Selective serotonin reuptake inhibitors.

### ***Table S5:* Risk factors for breakthrough infection in the post-booster cohort, stratified by levels of vigorous physical activity, probiotics, and angiotensin receptor blocker**

|  |  | **Stratified by vigorous physical exercise** | | **Stratified by probiotics** | | **Stratified by angiotensin receptor blockers** | |
| --- | --- | --- | --- | --- | --- | --- | --- |
|  |  | HR (95% CI) | p value | HR (95% CI) | p value | HR (95% CI) | p value |
|  | Age, years | 0.97 (0.96–0.98) | <0.0001 | 0.97 (0.96–0.98) | <0.0001 | 0.97 (0.96–0.98) | <0.0001 |
|  | Sex |  |  |  |  |  |  |
|  | Female | 1.00 |  | 1.00 |  | 1.00 |  |
|  | Male | 0.85 (0.73–0.99) | 0.0400 | 0.85 (0.73–0.99) | 0.0426 | 0.85 (0.73–0.99) | 0.0394 |
|  | Ethnicity |  |  |  |  |  |  |
|  | White | 1.00 |  | 1.00 |  | 1.00 |  |
|  | Mixed, multiple, or other ethnic groups | 0.28 (0.14–0.57) | 0.0004 | 0.28 (0.14–0.57) | 0.0004 | 0.28 (0.14–0.57) | 0.0004 |
|  | South Asian | 0.44 (0.21–0.94) | 0.0342 | 0.44 (0.21–0.92) | 0.0305 | 0.44 (0.21–0.92) | 0.0301 |
|  | Black, African, Caribbean, or Black British | 0.88 (0.33–2.37) | 0.7998 | 0.90 (0.33–2.41) | 0.8301 | 0.89 (0.33–2.40) | 0.8206 |
|  | Highest educational level attained |  |  |  |  |  |  |
|  | Post-grad | 1.00 |  | 1.00 |  | 1.00 |  |
|  | College or university | 1.07 (0.92–1.24) | 0.3927 | 1.06 (0.92–1.23) | 0.4054 | 1.06 (0.92–1.23) | 0.4071 |
|  | Higher or further (A levels) | 1.22 (1.00–1.49) | 0.0552 | 1.21 (0.99–1.48) | 0.0636 | 1.21 (0.99–1.48) | 0.0636 |
|  | Primary or secondary | 1.36 (1.08–1.72) | 0.0083 | 1.36 (1.08–1.71) | 0.0093 | 1.36 (1.08–1.71) | 0.0095 |
|  | Frontline worker |  |  |  |  |  |  |
|  | No | 1.00 |  | 1.00 |  | 1.00 |  |
|  | Non-health | 1.11 (0.90–1.36) | 0.3409 | 1.11 (0.90–1.37) | 0.3305 | 1.11 (0.90–1.36) | 0.3362 |
|  | Health or care | 0.91 (0.71–1.16) | 0.4394 | 0.91 (0.71–1.16) | 0.4485 | 0.91 (0.71–1.16) | 0.4488 |
|  | Housing |  |  |  |  |  |  |
|  | Owns own home | 1.00 |  | 1.00 |  | 1.00 |  |
|  | Mortgage | 1.19 (1.01–1.41) | 0.0407 | 1.19 (1.01–1.41) | 0.0396 | 1.20 (1.01–1.42) | 0.0371 |
|  | Privately renting | 1.39 (1.05–1.84) | 0.0205 | 1.38 (1.04–1.83) | 0.0246 | 1.38 (1.04–1.83) | 0.0237 |
|  | Renting from council | 1.07 (0.69–1.69) | 0.7527 | 1.08 (0.69–1.70) | 0.7253 | 1.09 (0.69–1.71) | 0.7110 |
|  | Other | 1.02 (0.67–1.57) | 0.9195 | 1.03 (0.67–1.58) | 0.8799 | 1.03 (0.67–1.59) | 0.8782 |
|  | Multigenerational households |  |  |  |  |  |  |
|  | Living alone | 0.92 (0.74–1.13) | 0.4262 | 0.92 (0.74–1.14) | 0.4338 | 0.92 (0.74–1.14) | 0.4332 |
|  | Single generation | 1.00 |  | 1.00 |  | 1.00 |  |
|  | Two or more generations | 0.98 (0.82–1.17) | 0.8120 | 0.98 (0.82–1.17) | 0.8125 | 0.98 (0.82–1.17) | 0.8046 |
|  | Shares home with schoolchildren (5–15 years) |  |  |  |  |  |  |
|  | No | 1.00 |  | 1.00 |  | 1.00 |  |
|  | Yes | 1.40 (1.09–1.79) | 0.0085 | 1.40 (1.09–1.79) | 0.0081 | 1.40 (1.09–1.79) | 0.0085 |
|  | Shares home with working-age adults (16–64 years) |  |  |  |  |  |  |
|  | No | 1.00 |  | 1.00 |  | 1.00 |  |
|  | Yes | 1.28 (1.08–1.52) | 0.0050 | 1.28 (1.08–1.52) | 0.0054 | 1.28 (1.08–1.52) | 0.0050 |
|  | Alcohol consumption, units per week |  |  |  |  |  |  |
|  | 0 | 1.00 |  | 1.00 |  | 1.00 |  |
|  | 1–7 | 1.06 (0.89–1.26) | 0.4982 | 1.06 (0.89–1.26) | 0.5020 | 1.06 (0.90–1.26) | 0.4879 |
|  | 8–14 | 1.02 (0.84–1.23) | 0.8656 | 1.01 (0.84–1.23) | 0.8926 | 1.01 (0.84–1.23) | 0.8807 |
|  | ≥15 | 1.31 (1.08–1.59) | 0.0058 | 1.30 (1.07–1.58) | 0.0073 | 1.31 (1.08–1.58) | 0.0063 |
|  | Travel outside of the UK since last questionnaire |  |  |  |  |  |  |
|  | No | 1.00 |  | 1.00 |  | 1.00 |  |
|  | Yes | 1.11 (0.94–1.30) | 0.2183 | 1.11 (0.94–1.30) | 0.2137 | 1.11 (0.94–1.30) | 0.2149 |
|  | Weekly journeys on public transport |  |  |  |  |  |  |
|  | 0 | 1.00 |  | 1.00 |  | 1.00 |  |
|  | 1–5 | 1.00 (0.85–1.18) | 0.9807 | 1.00 (0.85–1.17) | 0.9738 | 1.00 (0.85–1.17) | 0.9663 |
|  | ≥6 | 1.16 (0.92–1.46) | 0.2191 | 1.16 (0.92–1.46) | 0.2238 | 1.15 (0.91–1.45) | 0.2387 |
|  | Any visits to or from other households in past week |  |  |  |  |  |  |
|  | No | 1.00 |  | 1.00 |  | 1.00 |  |
|  | Yes | 1.10 (0.93–1.29) | 0.2772 | 1.10 (0.93–1.30) | 0.2734 | 1.09 (0.93–1.29) | 0.2876 |
|  | Weekly visits to shops |  |  |  |  |  |  |
|  | 0 | 1.00 |  | 1.00 |  | 1.00 |  |
|  | 1 | 0.93 (0.67–1.30) | 0.6756 | 0.94 (0.67–1.30) | 0.6950 | 0.94 (0.67–1.31) | 0.7114 |
|  | 2–3 | 1.06 (0.79–1.42) | 0.7101 | 1.06 (0.79–1.42) | 0.7098 | 1.07 (0.79–1.43) | 0.6700 |
|  | ≥4 | 1.17 (0.87–1.58) | 0.2951 | 1.17 (0.87–1.58) | 0.2948 | 1.18 (0.88–1.59) | 0.2739 |
|  | Weekly visits to other indoor public places (not shops) |  |  |  |  |  |  |
|  | None | 1.00 |  | 1.00 |  | 1.00 |  |
|  | 1–2 | 1.09 (0.91–1.31) | 0.3301 | 1.10 (0.92–1.31) | 0.3186 | 1.09 (0.91–1.31) | 0.3251 |
|  | ≥3 | 1.32 (1.10–1.60) | 0.0036 | 1.33 (1.10–1.60) | 0.0036 | 1.33 (1.10–1.61) | 0.0033 |
|  | Vigorous physical exercise, h per week |  |  |  |  |  |  |
|  | 0 | .. | .. | 0.89 (0.75–1.06) | 0.1843 | 0.89 (0.75–1.06) | 0.1826 |
|  | 1 | .. | .. | 0.91 (0.78–1.07) | 0.2642 | 0.91 (0.78–1.07) | 0.2539 |
|  | 2 | .. |  | 1.00 |  | 1.00 |  |
|  | Actual sleep, h/night |  |  |  |  |  |  |
|  | ≤5 | 0.73 (0.57–0.93) | 0.0100 | 0.73 (0.57–0.93) | 0.0100 | 0.73 (0.57–0.93) | 0.0102 |
|  | 6 | 1.02 (0.87–1.18) | 0.8398 | 1.01 (0.87–1.18) | 0.8664 | 1.02 (0.88–1.18) | 0.8315 |
|  | 7 | 1.00 |  | 1.00 |  | 1.00 |  |
|  | ≥8 | 1.01 (0.85–1.20) | 0.8846 | 1.01 (0.85–1.20) | 0.8875 | 1.01 (0.85–1.20) | 0.8931 |
|  | Weekly SARS-CoV-2 incidence | 1.09 (1.08–1.10) | <0.0001 | 1.09 (1.08–1.10) | <0.0001 | 1.09 (1.08–1.10) | <0.0001 |
|  | Inter-vaccine interval (primary to booster), weeks | 0.98 (0.96–1.01) | 0.1794 | 0.98 (0.96–1.01) | 0.1876 | 0.98 (0.96–1.01) | 0.1914 |
|  | Combination of primary and booster vaccinations |  |  |  |  |  |  |
|  | ChAdOx1 plus BNT162b2 booster | 1.06 (0.91–1.23) | 0.4630 | 1.05 (0.91–1.23) | 0.4991 | 1.05 (0.91–1.23) | 0.5006 |
|  | ChAdOx1 plus mRNA-1273 booster | 1.29 (1.03–1.61) | 0.0279 | 1.28 (1.03–1.61) | 0.0291 | 1.28 (1.02–1.60) | 0.0315 |
|  | BNT162b2 plus BNT162b2 booster | 1.00 |  | 1.00 |  | 1.00 |  |
|  | BNT162b2 plus mRNA-1273 booster | 1.00 (0.68–1.47) | 0.9852 | 1.00 (0.68–1.47) | 0.9836 | 0.99 (0.67–1.46) | 0.9673 |
|  | Season of booster vaccination |  |  |  |  |  |  |
|  | Mid-October to mid-April (Winter) | 1.00 |  | 1.00 |  | 1.00 |  |
|  | Mid-April to mid-October (Summer) | 0.55 (0.45–0.67) | <0.0001 | 0.55 (0.45–0.67) | <0.0001 | 0.55 (0.46–0.67) | <0.0001 |
|  | Previous infection |  |  |  |  |  |  |
|  | No evidence of previous infection | 1.00 |  | 1.00 |  | 1.00 |  |
|  | Previous infection before primary course of vaccination | 0.77 (0.57–1.04) | 0.0892 | 0.77 (0.56–1.04) | 0.0857 | 0.76 (0.56–1.03) | 0.0816 |
|  | Previous infection after primary course of vaccination | 0.26 (0.15–0.45) | <0.0001 | 0.26 (0.15–0.45) | <0.0001 | 0.26 (0.15–0.45) | <0.0001 |
|  | Probiotics |  |  |  |  |  |  |
|  | No | 1.00 |  | .. |  | 1.00 |  |
|  | Yes | 0.80 (0.58–1.10) | 0.1749 | .. | .. | 0.80 (0.58–1.11) | 0.1806 |
|  | Fish oil, krill oil, or other omega-3 supplements |  |  |  |  |  |  |
|  | No | 1.00 |  | 1.00 |  | 1.00 |  |
|  | Yes | 0.83 (0.66–1.04) | 0.1032 | 0.83 (0.66–1.04) | 0.1039 | 0.83 (0.66–1.04) | 0.1007 |
|  | Cod liver oil supplements |  |  |  |  |  |  |
|  | No | 1.00 |  | 1.00 |  | 1.00 |  |
|  | Yes | 0.80 (0.61–1.04) | 0.1004 | 0.79 (0.61–1.04) | 0.0936 | 0.79 (0.60–1.04) | 0.0904 |
|  | Atopy |  |  |  |  |  |  |
|  | No | 1.00 |  | 1.00 |  | 1.00 |  |
|  | Yes | 0.90 (0.78–1.05) | 0.1723 | 0.91 (0.78–1.05) | 0.1848 | 0.90 (0.78–1.04) | 0.1695 |
|  | Diabetes types |  |  |  |  |  |  |
|  | No diabetes | 1.00 |  | 1.00 |  | 1.00 |  |
|  | Pre-diabetes | 0.87 (0.57–1.34) | 0.5346 | 0.87 (0.57–1.33) | 0.5175 | 0.87 (0.57–1.33) | 0.5191 |
|  | Type 1 diabetes | 0.88 (0.44–1.78) | 0.7242 | 0.89 (0.44–1.79) | 0.7391 | 0.88 (0.44–1.78) | 0.7300 |
|  | Type 2 diabetes | 0.91 (0.53–1.57) | 0.7384 | 0.91 (0.53–1.57) | 0.7442 | 0.91 (0.53–1.57) | 0.7385 |
|  | Beta blockers |  |  |  |  |  |  |
|  | No | 1.00 |  | 1.00 |  | 1.00 |  |
|  | Yes | 0.86 (0.65–1.14) | 0.3017 | 0.86 (0.65–1.15) | 0.3083 | 0.86 (0.65–1.15) | 0.3134 |
|  | ACE inhibitors |  |  |  |  |  |  |
|  | No | 1.00 |  | 1.00 |  | 1.00 |  |
|  | Yes | 1.01 (0.79–1.29) | 0.9364 | 1.01 (0.79–1.29) | 0.9393 | 1.01 (0.79–1.29) | 0.9351 |
|  | Angiotensin receptor blockers |  |  |  |  |  |  |
|  | No | 1.00 |  | 1.00 |  | .. |  |
|  | Yes | 0.91 (0.67–1.23) | 0.5446 | 0.91 (0.67–1.23) | 0.5397 | .. | .. |
|  | Thiazides |  |  |  |  |  |  |
|  | No | 1.00 |  | 1.00 |  | 1.00 |  |
|  | Yes | 0.60 (0.36–1.00) | 0.0480 | 0.60 (0.36–0.99) | 0.0450 | 0.60 (0.36–0.99) | 0.0462 |
|  | Metformin |  |  |  |  |  |  |
|  | No | 1.00 |  | 1.00 |  | 1.00 |  |
|  | Yes | 0.83 (0.42–1.63) | 0.5813 | 0.83 (0.42–1.63) | 0.5830 | 0.82 (0.41–1.62) | 0.5670 |

ACE = angiotensin-converting-enzyme. ChAdOx1 = ChAdOx1 nCoV-19. HR = hazard ratio.
